# A comprehensive antigen production and characterization study for easy-to-implement, highly specific and quantitative SARS-CoV-2 antibody assays

**DOI:** 10.1101/2021.01.19.21249921

**Authors:** Miriam Klausberger, Mark Dürkop, Helmuth Haslacher, Gordana Wozniak-Knopp, Monika Cserjan-Puschmann, Thomas Perkmann, Nico Lingg, Patricia Pereira Aguilar, Elisabeth Laurent, Jelle De Vos, Manuela Hofer, Barbara Holzer, Maria Stadler, Gabriele Manhart, Klemens Vierlinger, Margot Egger, Lisa Milchram, Elisabeth Gludovacz, Nicolas Marx, Christoph Köppl, Christopher Tauer, Jürgen Beck, Daniel Maresch, Clemens Grünwald-Gruber, Florian Strobl, Peter Satzer, Gerhard Stadlmayr, Ulrike Vavra, Jasmin Huber, Markus Wahrmann, Farsad Eskandary, Marie-Kathrin Breyer, Daniela Sieghart, Peter Quehenberger, Gerda Leitner, Robert Strassl, Alexander E. Egger, Christian Irsara, Andrea Griesmacher, Gregor Hoermann, Günter Weiss, Rosa Bellmann-Weiler, Judith Loeffler-Ragg, Nicole Borth, Richard Strasser, Alois Jungbauer, Rainer Hahn, Jürgen Mairhofer, Boris Hartmann, Nikolaus B. Binder, Gerald Striedner, Lukas Mach, Andreas Weinhäusl, Benjamin Dieplinger, Florian Grebien, Wilhelm Gerner, Christoph J. Binder, Reingard Grabherr

**Affiliations:** Department of Biotechnology, University of Natural Resources and Life Sciences (BOKU) Vienna, Austria; Novasign GmbH Vienna, Austria; Department of Laboratory Medicine, Medical University of Vienna, Austria; CD Laboratory for innovative Immunotherapeutics, Vienna, Austria; ACIB-Austrian Centre of Industrial Biotechnology, Graz, Austria; BOKU Core Facility Biomolecular & Cellular Analysis, University of Natural Resources and Life Sciences Vienna, Austria; Department of Chemical Engineering, Vrije Universiteit Brussel (VUB), Brussels, Belgium; Competence Unit Molecular Diagnostics, Center for Health and Bioresources, AIT Austrian Institute of Technology GmbH, Vienna, Austria; Austrian Agency for Health and Food Safety (AGES), Department for Animal Health, Moedling, Austria; Institute of Immunology, University of Veterinary Medicine Vienna, Austria; Institute for Medical Biochemistry, University of Veterinary Medicine, Vienna, Austria; Department of Laboratory Medicine, Konventhospital Barmherzige Brueder Linz and Ordensklinikum Linz Barmherzige Schwestern, Linz, Austria; BOKU Core Facility Mass Spectrometry, University of Natural Resources and Life Sciences, Vienna, Austria; Department of Chemistry (BOKU), University of Natural Resources and Life Sciences Vienna, Austria; enGenes Biotech GmbH, Vienna, Austria; Department of Applied Genetics and Cell Biology (DAGZ), University of Natural Resources and Life Sciences (BOKU) Vienna, Austria; Department of Medicine III, Division of Nephrology and Dialysis, Medical University Vienna, Austria; Department of Respiratory and Critical Care Medicine and Ludwig Boltzmann Institute for Lung Health, Otto Wagner Hospital, Vienna, Austria; Division of Rheumatology, Department of Medicine III, Medical University of Vienna, Austria; Department of Blood Group Serology and Transfusion Medicine, Medical University of Vienna, Austria; Central Institute for Medical and Chemical Laboratory Diagnosis, Innsbruck University Hospital, Innsbruck, Austria; MLL Munich Leukemia Laboratory, Munich, Germany; Department of Internal Medicine II, Innsbruck Medical University, Austria; Technoclone Herstellung von Diagnostika und Arzneimitteln GmbH, Vienna, Austria; Christian Doppler Laboratory for an Optimized Prediction of Vaccination Success in Pigs, University of Veterinary Medicine, Vienna, Austria; The Pirbright Institute, Pirbright, United Kingdom

## Abstract

Antibody tests are essential tools to investigate humoral immunity following SARS-CoV-2 infection. While first-generation antibody tests have primarily provided qualitative results with low specificity, accurate seroprevalence studies and tracking of antibody levels over time require highly specific, sensitive and quantitative test setups. Here, we describe two quantitative ELISA antibody tests based on the SARS-CoV-2 spike receptor-binding domain and the nucleocapsid protein. Comparative expression in bacterial, insect, mammalian and plant-based platforms enabled the identification of new antigen designs with superior quality and high suitability as diagnostic reagents. Both tests scored excellently in clinical validations with multi-centric specificity and sensitivity cohorts and showed unprecedented correlation with SARS-CoV-2 neutralization titers. Orthogonal testing increased assay specificity to 99.8%, thereby enabling robust serodiagnosis in low-prevalence settings. The inclusion of a calibrator permits accurate quantitative monitoring of antibody concentrations in samples collected at different time points during the acute and convalescent phase of COVID-19.

## 1. Introduction

Serological testing of severe acute respiratory syndrome coronavirus 2 (SARS-CoV-2) infections remains an essential tool for seroprevalence studies and complements PCR-based diagnosis in identifying asymptomatic individuals (*1*). Antibody tests are gaining additional importance as means to characterize the extent of infection- or vaccine-induced immunity. To cope with the urgent demand for sensitive and reliable test systems, many manual and automated serological tests for coronavirus disease 2019 (COVID-19) became available within a short period of time (*2*). Owing to the acuity of a spreading pandemic, many of these early developed test systems lacked adequate validation and thereby fueled mistrust, while stocks of others were exhausted rapidly due to increased demand (*3*).

Antigen selection and quality are crucial aspects of assay development and influence diagnostic performance (*4*), such as sensitivity and specificity as well as assay availability, scalability and their field of application. Ideal candidate antigens for *in-vitro* serodiagnosis are highly immunogenic, support early and robust detection of seroconversion after an infection and result in low false positivity rates. Additionally, production platforms supporting high process yields ensure sustainable assay supply. To date, biotechnological performance attributes and their influence on serodiagnostics were not reported during the development of assays for SARS-CoV-2 detection. Likewise, no comprehensive study comparing and validating the same SARS-CoV-2 antigen produced in different expression systems with larger cohorts is available.

In this study, we developed two quantitative, enzyme-linked immunosorbent assay (ELISA)-based serotests relying on SARS-CoV-2 receptor-binding domain (RBD) and nucleocapsid protein (NP) antigens of superior design and quality. Since these assays utilize established ELISA technology, they are easy to implement in any lab worldwide. We describe a comprehensive approach assessing biotechnological parameters such as antigen quality attributes and manufacturability for an ideal test setup. For this purpose, we compared several animal cell lines and plant-based expression platforms for their ability to support high-quantity and quality RBD production and assessed whether the employed production host influences antigen performance. We extensively validated the tests for clinical utility featuring sera from individuals covering the full spectrum of disease presentations at different time points post infection and a large specificity cohort including samples with antibodies towards human coronaviruses (hCoVs) and those from individuals with underlying non-infectious diseases. Moreover, we validated the tests with time-resolved acute and early convalescent samples from hospitalized patients and showed that only RBD-specific antibodies demonstrate excellent correlation with neutralization assays already in the early phase of infection.

## 2. Results

### Comparative profiling of expression hosts for SARS CoV-2 RBD and NP production for diagnostic use

Initially, five eukaryotic expression systems were compared for their capacity to support high-quantity and high-quality expression of the glycosylated SARS-CoV-2 spike RBD. Our pre-defined quality attributes covered activity in a functional binding assay using a conformation-dependent RBD-specific antibody (CR3022), protein integrity and glycosylation determined by mass spectrometry, as well as manufacturability (**Fig. 1A**). Biolayer interferometry analysis revealed that RBD obtained from different mammalian and insect expression systems have comparable affinities (range: 21 – 43 nM) for the mAb CR3022 (**Fig. 1A, left panel**). Glycan analysis confirmed host-specific N-glycosylation of the respective proteins, which was of complex-type for the human (HEK-6E) and non-human mammalian cell lines (CHO-K1, CHO-S) as well as for plant (*Nicotiana benthamiana*)-derived RBD. We found paucimannosidic N-glycosylation for the *Trichoplusia ni* insect cell line (*Tnms*42)-derived RBD (**Fig. S1A, B**). Peptide mapping verified the integrity of the protein primary structure (data not shown). Unoptimized and small-scale electroporation of non-human (CHO-K1 and CHO-S) and baculovirus infection of insect (*Tnms*42) cell lines produced overall yields after purification of less than one mg RBD per liter of culture. Polyethylenimine (PEI) transfection of HEK cells readily provided higher overall volumetric yields (∼40 mg/L) without further process optimization (**Fig. 1A, left panel**). Analytical size-exclusion chromatography (HP-SEC) revealed expression platform and production batch-dependent RBD homodimer contents. (**Fig. S2**). For plant-expressed RBD, dimerization was particularly pronounced. We identified an unpaired cysteine residue (Cys538) close to the C-terminus of the canonical RBD sequence as a possible cause for RBD dimerization. A truncated RBD construct (tRBD) lacking this cysteine residue was less prone to homodimer formation, but retained full functionality in the binding assay and similar expression yields (**Fig. 1A, Fig. S3**). From a manufacturing perspective, tRBD thus provided less batch-to-batch variation, which is a pre-requisite for a diagnostic antigen.

**Fig. 1.**
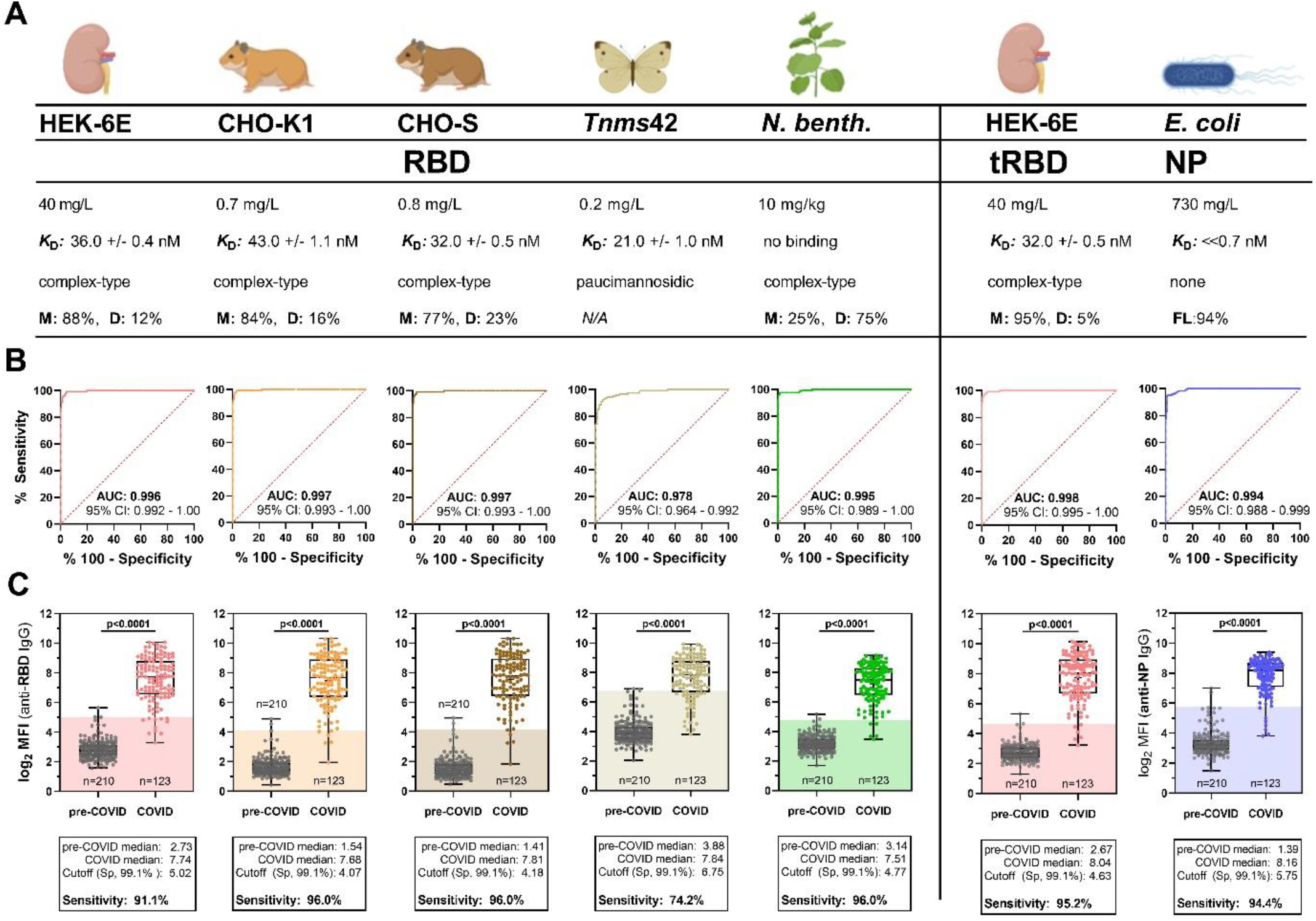
Comparative profiling of SARS CoV-2 antigens from different expression hosts for serodiagnosis. **A-C**, the canonical SARS-CoV-2 RBD expressed in five biotechnological platforms (HEK, CHO-K1, CHO-S, *Tnms*42, *N. benthamiana*, **left panel**), an optimized RBD construct expressed in HEK cells (tRBD) as well as the NP produced in *E. coli* **(right panel)** were compared in terms of biotechnological parameters as well as seroreactivity to identify ideal candidates that may be sustainably produced for specific and sensitive SARS-CoV-2 serodiagnosis. **A)** Pre-defined process and protein quality parameters include overall yield after purification, functional binding to the conformation-dependent mAb CR3022 (RBD) or a commercially available anti-NP antibody as verified by biolayer interferometry, as well as glycosylation analysis. Purified monomer (M), dimer (D), and NP full-length protein (FL)-content was determined by HP-SEC **B-C**, Pre-validation of antigens for serodiagnosis with sera of healthy blood donors collected prior to 2018 (n=210) and convalescent sera from a COVID cohort (n=124; see methods for cohort description) with an automatable Luminex bead-based serotest. **B)** Receiver operating characteristic (ROC) curves of the assayed antigens with an indication of the area under the curve (AUC) and 95% confidence interval (CI), **C)** Seroreactivity of the two cohorts at a final serum dilution of 1:1,200. Blank-corrected values are shown. Shades indicate the calculated cut-off yielding a specificity (Sp) of 99.1% for comparison of antigen performance. P-values were calculated by Mann-Whitney U tests.

To assess the performance of the antigens for discrimination between sera from SARS-CoV-2-exposed (n=124) and uninfected individuals (n=210), we applied a high-throughput (HTP) automated bead-based multiplex assay (**Fig. 1B, C**). The performance of diagnostic tests are commonly assessed through receiver operating characteristic (ROC) curves and the analysis of area under the ROC curve (AUC-ROC). ROC curves are simple graphical representations of the relationship between sensitivity and specificity of a test over all possible diagnostic cut-off values and AUCs give the overall ability of a test to discriminate between two populations (*5*). We used theses analyses to assess potential differences in the diagnostic performance of RBD from different expression hosts. Almost all antigens at this high purity demonstrated AUC values of >0.99, demonstrating the high suitability of the RBD from any source as diagnostic antigen. The AUC value of insect-derived RBD was slightly lower (AUC: 0.978 [0.964-0.992]); the differences, however, were not significant (**Fig. 1B**). We then applied antigen-specific cut-offs to compare the performance of the antigens at a pre-defined consensus specificity of 99.1%. At this criterion, we obtained high sensitivities (range 94.4%-96.0%) with all antigens, except for insect-derived RBD. There, seroreactivity with pre-COVID-19 sera was about 2^2^- (4)-fold higher than observed for CHO-expressed RBDs. This resulted in 26% of COVID-19 sera to fall below the threshold, increasing the rate of false-negatives **(Fig. 1C)**. The tRBD displayed a comparable seroreactivity profile to the RBD.

During our pre-validation experiments we observed a strong effect of residual host cell proteins on assay performance (**Fig. S4**), even in formulations derived from human cell lines. Therefore, RBD/tRBDs were purified via an Immobilized metal affinity chromatography (IMAC) capture followed by a scalable and fast flow-through Anion exchange (AIEX) chromatography step, leading to purities of up to 99%. Owing to reproducible highest production yields of functional protein with adequate diagnostic performance and less batch-to-batch variation, we decided to pursue with HEK-expressed tRBD for our further validations.

As the NP of SARS-CoV-1 has been described to be well produced in bacteria (*6*), we decided to produce the SARS-CoV-2 NP in *Escherichia coli*. We combined two recently developed generic manufacturing strategies, the CASPON (cpCasp2-based platform fusion protein process) technology (*7*) and the enGenes-X-press technology (*8*), allowing for high-level soluble expression of heterologous proteins. NP was expressed as a fusion protein with an N-terminally fused CASPON tag that enables affinity purification and can afterwards be efficiently proteolytically removed, thereby generating the authentic N-terminus. High soluble volumetric titers of 3.7 g/L in a growth-decoupled fed-batch production process yielded 730 mg/L NP after purification with a modified CASPON platform process (**Fig. 1A, right panel**). This strategy delivered untagged NP protein at exceptionally high quality (93.6% purity, defined as protein full-length content) after a multi-step-downstream process. The remaining impurities consisted of NP-related fragments and RNA. Host cell protein concentration was 0.9 ng/mg NP and dsDNA concentration was 1 µg/mg NP, as determined by De Vos and colleagues (*9*). NP has an intrinsic propensity to oligomerize and displays very slow dissociation from the antibody (Abcam, ab272852). Therefore, we provide an upper limit for the *K*_D_ value, and calculated *k*_obs_ values as a surrogate kinetic parameter instead (**Fig. 1A, right panel, Fig. S3**). The nucleocapsid protein also presented with excellent AUC values of 0.994 (0.988-0.999) and comparable performance to HEK-derived tRBD. While the seroreactivity profile of pre-COVID sera appeared to be more heterogenous against the NP than for tRBD, COVID sera demonstrated a more consistent, robust response against the nucleocapsid protein (**Fig 1B, C, right panel**). This comprehensive set of biotechnological and assay performance characteristics prompted us to pursue ELISA test development with HEK-expressed tRBD and bacterially produced NP.

### Assessment of antigen-dependency of false-positive and false-negative results

A set of sera (28-31 convalescence sera from the above tested), that was considered to be particularly challenging since it included 80% of the identified outliers or borderline serum samples, was selected to optimize the conditions for an ELISA with tRBD and NP. At the final conditions (6 µg/mL coating antigen, 1:200 serum dilution) we achieved high sensitivities of 85.7% (Luminex) or 100% (ELISA) at the pre-defined consensus specificity criteria (99.1%, Luminex, 92.9% ELISA, **Fig. 2A**) with both antigens. Our findings from the Luminex antigen pre-validations were in good agreement with the ELISA results, as was demonstrated by the excellent cross-platform correlation of the seroreactivity readouts (tRBD: r_s_=0.97, p<0.0001; NP: r_s_=0.87, p <0.0001,). Next, we aimed to assess whether false-positive or false-negative results are independent of the test antigen. With both test formats, up to 50% of the false-negative samples did not simultaneously react with both antigens (**Fig. 2B**). Concurrently, none of the false-positive sera in the ELISA, and only 20% of the false-positive sera (5 out of 25) in the Luminex test simultaneously reacted with both the tRBD and NP (**Fig. 2C**). Levels of tRBD- and NP-specific antibodies correlated well with each other (r_s_=0.75-0.80, p<0.0001, **Fig. 2B**) and also with the ability of the respective sera to neutralize authentic SARS-CoV-2 virus. Yet, with partial correlation analysis we could demonstrate that only anti-tRBD antibodies do have a causal relation with viral neutralization (r_s_=0.68, p=0.0003, **Fig. 2D**).

**Fig. 2.**
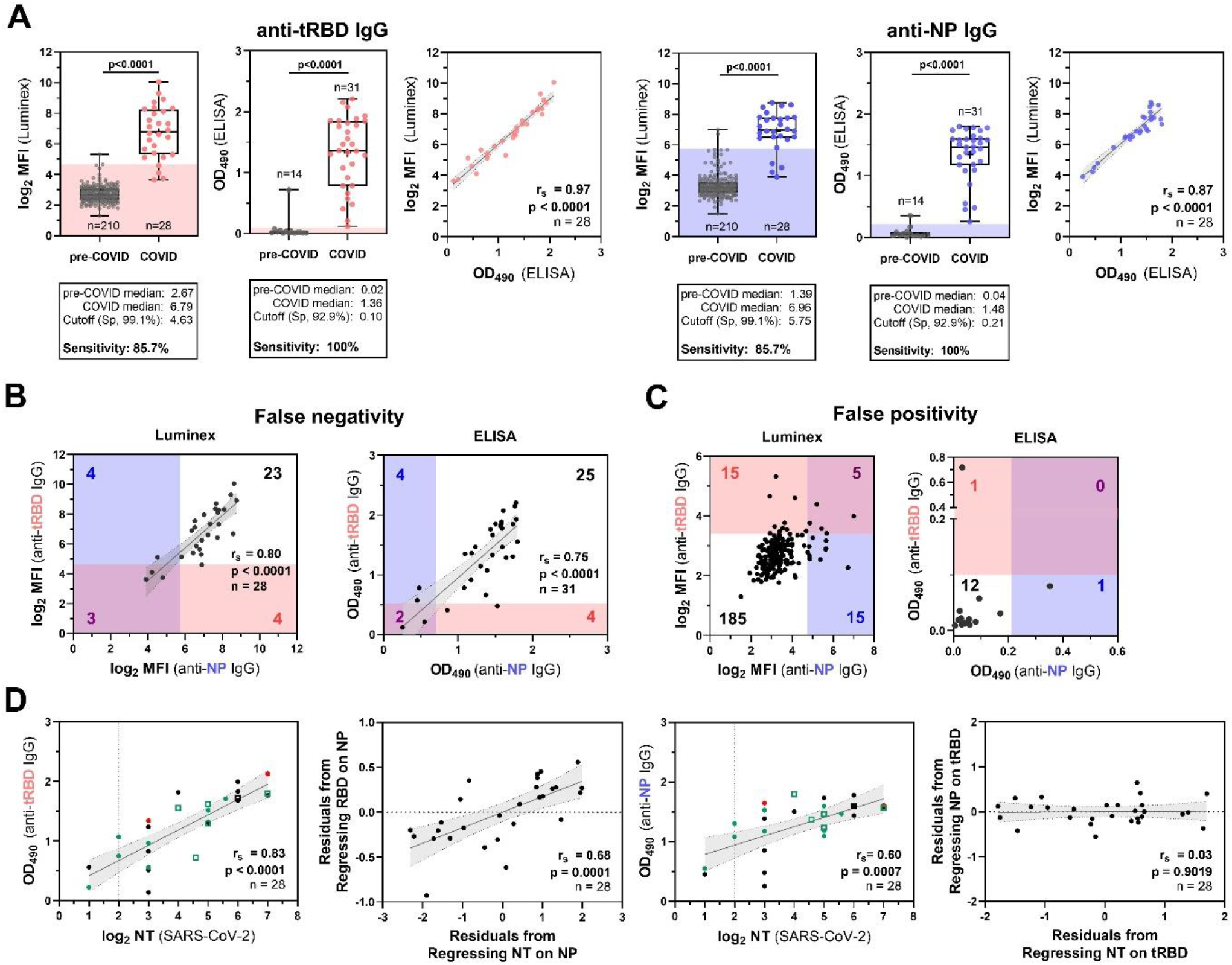
Convalescent sera from blood donors with mild to moderate courses of disease indicate an advantage of dual-antigen testing and a correlation of tRBD-specific antibodies with SARS-CoV-2 neutralization. **A-D**, A small set of convalescent sera (n=28-31, part of the Medical University of Vienna COVID-19-cohort) with described courses of disease was used for in-depth analysis of the ELISA candidate antigens. Pre-COVID-19 sera included blood donor sera (n=210 and n=14) collected in pre-COVID-19 times (see methods for detailed cohort description). **A)** Seroreactivity of HEK-tRBD and *E. coli*-derived NP as assessed by the Luminex platform and ELISA at serum dilutions of 1:1,200 and 1:200, respectively, and the cross-platform correlation of the respective readouts. Data give the mean of blank-corrected values from three independent production batches. Sensitivities with the respective test antigens at the indicated pre-defined specificities were calculated by AUC-analysis of ROC curves, P-values were calculated by Mann-Whitney U tests. **B-C**, Assessment of overlaps in **B)** false-negative and **C)** false-positive serum samples identified with both the tRBD or NP antigen in the Luminex and ELISA assay. The cut-offs were set to yield low sensitivity (87.1%, ELISA; 85.,7%, Luminex) or specificity (92.9%, both assays), respectively. Shades are colored according to the respective antigens (NP: blue, tRBD: pink) and, and indicate the cut-offs. Numbers in blue and red give the total numbers of false-positives/false-negatives for NP or tRBD, respectively, while purple numbers give false-positives/negatives identified with both antigens. **D)** Correlation and partial correlation analysis of ELISA anti-tRBD as well as anti-NP levels with neutralization titers obtained with authentic SARS-CoV-2 virus. Partial correlations take the effect of antibody levels towards the respective other antigen into account Individual sera are color-coded according to the course of disease (green: asymptomatic and mild; black: moderate; red: severe). Solid lines indicate the linear regression and shades with dotted borders give the 95% CI. Full circles are for sera from individuals with a PCR-confirmed SARS-CoV-2 infection, open squares indicate asymptomatic close contacts. r_s_, Spearman’s correlation factor.

### Cut-off modeling and diagnostic performance of the tests in a large validation cohort

The above data provided an indication that reactivity of COVID-19 sera is dependent on the test antigen, fostering the idea for combined use in applications requiring high specificity. Test kits for both antigens were generated (termed Technozym NP or RBD IgG Test, Technoclone, Vienna, AT), providing the antigens in lyophilized form at a coating concentration of 6µg/mL. The kits included a five-point calibrator set, based on the RBD-specific antibody CR3022, to enable quantitative readouts and further expand the tests’ application fields.

Both the tRBD and the NP ELISA were evaluated using 244 samples from patients with active or previous SARS-CoV-2 infection covering the full spectrum of disease presentations (asymptomatic to individuals requiring intensive care). The large specificity cohort (n=1,126) covered a great variety in samples from pre-COVID times including sera from individuals with rheumatic disease, human coronavirus infections drawn during winter months to increase the likelihood for respiratory infections. A detailed description of the SARS-CoV-2 positive cohorts can be found in **Table S1**. In ROC-analysis, both assays presented with excellent areas under the curve (tRBD: 0.976, NP: 0.974, **Fig. 3A, B)**. The Youden index was maximal at a cut-off of >2.549 U/mL for tRBD (Youden index=0.901) and at >3.010 U/mL for NP (Youden index=0.882) yielding high sensitivities (tRBD: 95.8% [91.6-97.4], NP: 93.0% [89.1-95.9] at these cut-offs. Yet, specificities (tRBD 95.3% [93.6-96.2], NP 95.1% [93.7-96.3]) were insufficient to yield satisfactory positive predictive values (PPVs), which give the probabilities that an individual with a positive test result indeed has antibodies for SARS-CoV-2. At a low seroprevalence rate of 5% the PPVs at these cut-offs would be equivalent to a coin toss, with 50.2% (43.8-56.5) for tRBD and 50.1% (43.6-56.5) for NP. To increase assay specificity of each test individually, thereby increasing predictability at low seroprevalences, cut-off criteria based on the 99^th^ percentile method were established. Ninety-nine percent of all negative samples showed results below 7.351 (95% CI: 5.733-10.900) U/mL for the tRBD and 7.748 (5.304-11.157) U/mL for the NP ELISA. When shifting the cut-off to 8.000 U/mL (taking a safety margin into account), specificities increased to 99.2% for the tRBD and 99.1% for the NP ELISA. At the same time, sensitivities slightly dropped to 86.3% and 76.7% for the tRBD and NP assays, respectively. The PPVs increased to 84.8% for tRBD and 82.5% for NP **(Fig. 3A, B)**. To monitor of immune responses after infection or vaccination, a cut-off yielding higher sensitivities at acceptable specificities was established. A cut-off between the criteria suggested by the ROC analysis and that calculated by the 99^th^ percentile method, e.g., 5.000 U/mL, yielded a sensitivity of 89.8% and a specificity of 98.0% for the tRBD assay, as well as a sensitivity of 86.5% and a specificity of 98.3% for the NP assay **(Fig. 3A, B)**.

**Fig. 3.**
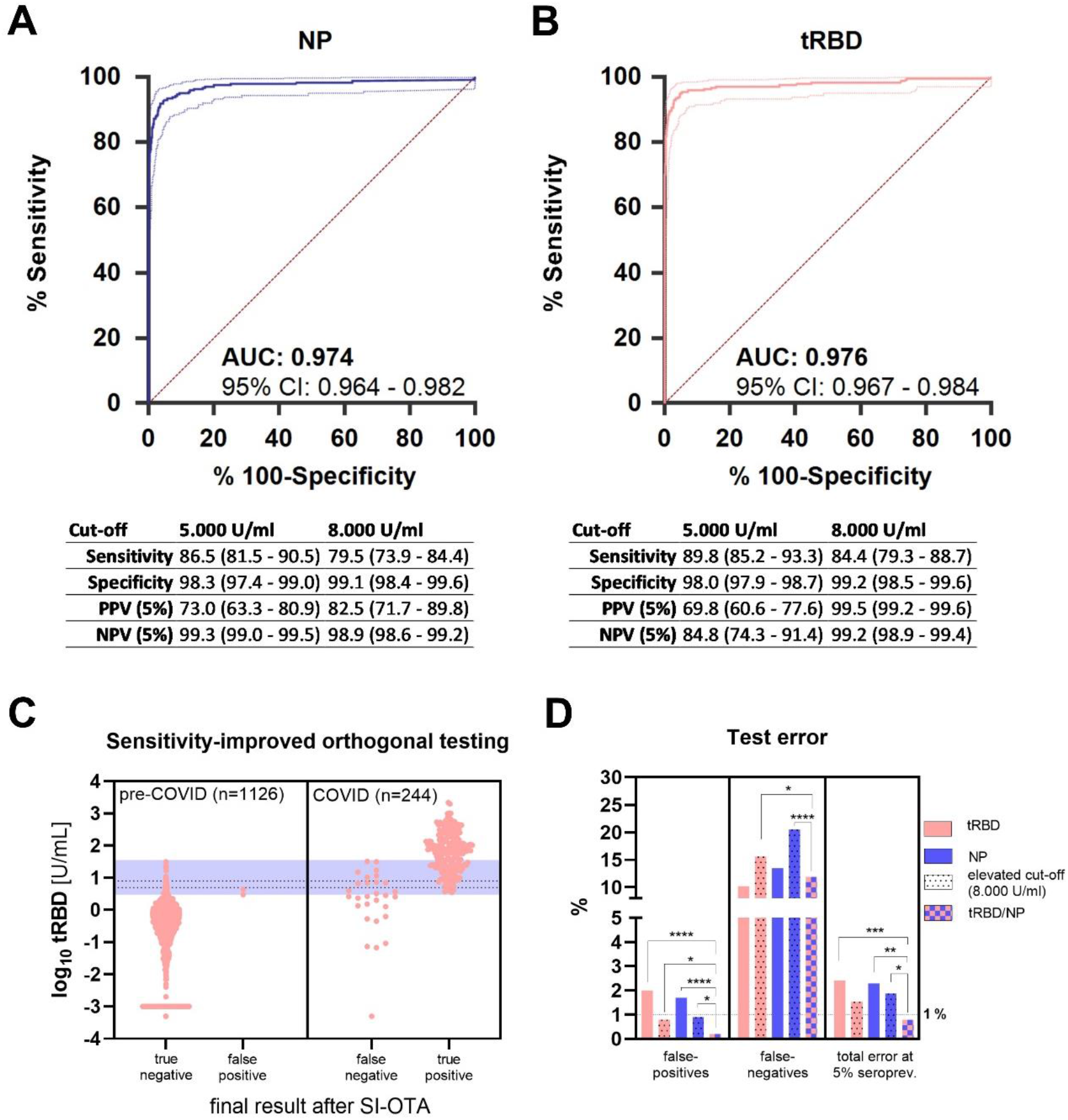
Performance validation of the Technozym NP and RBD tests. ROC-curve (AUC±95% confidence intervals) of **A)** the Technozym RBD- and **B)** the NP-ELISA on basis of a cohort of 1,126 pre-COVID-19 and 244 COVID-19 serum samples. **C)** Results from an adaptive orthogonal testing approach, where all samples yielding <3.000 U/mL in the tRBD ELISA were considered negative and samples with tRBD >35.000 U/mL positive. Samples with tRBD values between those borders were re-tested with the NP ELISA (blue shade). If NP>3.500 U/mL, positivity was confirmed, otherwise it was ruled out. Dashed lines indicate the cut-offs determined by the 99^th^ percentile method (8.000 U/mL) and a reduced cut-off with increased sensitivity (5.000 U/mL, between 99^th^ percentile- and Youden-index criteria) to display the increase in sensitivity gained by the orthogonal test system. **D)** Differences in false-positive and -negative test results for different individual and combined test setups were compared by z-tests, total errors at an estimated 5% seroprevalence were compared by χ^2^-tests for proportions. ∗ *P*<0.05, ∗∗ *P*<0.01, ∗∗∗ *P*<0.001, ∗∗∗∗ *P*<0.0001.

### Orthogonal testing approach at very low seroprevalences to approximate 100% specificity

For low seroprevalences, when specificities need to approximate 100% in order to achieve acceptable PPVs, we considered an orthogonal testing approach (OTA). Our previous experiments already provided an indication that false-positives among pre-COVID-19 sera do not necessarily react with both antigens (**Fig. 2C**). As a classical OTA might negatively affect sensitivities an adaptive, sensitivity-improved (SI-OTA) was applied (*10*). To this end, the above-described validation cohorts were screened with the tRBD ELISA. All samples with results ranging between the cut-off defined by the Youden index (taking a safety margin of, *i*.*e*., 3.000 U/mL into account) and 35.000 U/mL (as no false-positives occurred above 31.500 U/mL) were re-tested with the NP ELISA. There, also the Youden index criterion, adding a safety margin, was applied for positivity (>3.500 U/mL). Samples with <3.000 U/mL in the screening test were considered negative; samples with results between 3.000 U/mL and 35.000 U/mL in the screening tests and at the same time >3.500 U/mL in the confirmation test were considered positive; samples >35.000 U/mL in the screening test were considered positive. Applying these criterions 133 of 1,370 samples needed to be re-tested. In turn, this approach led to a significantly enhanced specificity (99.8% [99.4-100.0]) when compared to the tRBD test alone both at a cut-off of 5.000 U/mL (+0.019, P<0.0001) and 8.000 U/mL (+0.006, P=0.035). Compared to the latter, sensitivity (88.1% [83.4-91.9]) was improved (+0.037, p<0.050) and the PPV rose to 96.3% (86.7-99.1), see **Fig. 3C**. To achieve this improvement, only 133 (*i*.*e*., those with tRBD levels between 3.000 and 35.000 U/mL) of the overall 1,370 samples needed to be re-tested by the NP assay, resulting in less than 10% increase in testing volume.

### Cross-reactivity of SARS-CoV-2 IgG antibodies with endemic and seasonal coronaviruses

To better characterize our specificity cohorts, we explored the prevalence of antibodies towards common cold coronaviruses and possible cross-reactivities with our assays. To do so, outliers among the pre-COVID-19 cohort were defined as sera with readouts higher than the 75^th^ percentile + 1.5x interquartile range (IQR) of the total cohort seroreactivity towards the SARS-CoV-2 NP or tRBD (outlier NP: n=17; tRBD: n=4). Above these cutoffs, all sera from our specificity cohorts reacted strongly with the spike proteins of circulating human coronaviruses (hCoVs) HKU-1, OC43, 229E, and NL63, confirming widespread seroprevalence in the general population (**Fig. 4A, B)**. To further characterize the identified outliers among the pre-COVID-19 sera, we calculated their relative IgG signals, set them in relation to a roughly equal number of sera located at the other extreme on the seroreactivity scale (sera with readouts <25^th^ percentile toward the respective antigen) and compared the differences in relative IgG levels to that towards hCoV antigens. Among our pre-validation cohort, sera with highest relative reactivity towards NP (mean difference: 0.88, p>0.0001) also demonstrated significantly elevated relative median IgG levels towards the spike protein of HKU-1 (mean difference: 0.13, p=0.0113, **Fig. 4B**). The specificity cohort we used for clinical validation included 8 sera from individuals with PCR-confirmed hCoV infection. None of these yielded false-positive readouts at a cutoff of 5.000 U/mL (**Fig. 4C**) at comparably low specificities of 95.3% (tRBD) and 96.1% (NP) (see **Fig. 3A, B**).

**Fig. 4.**
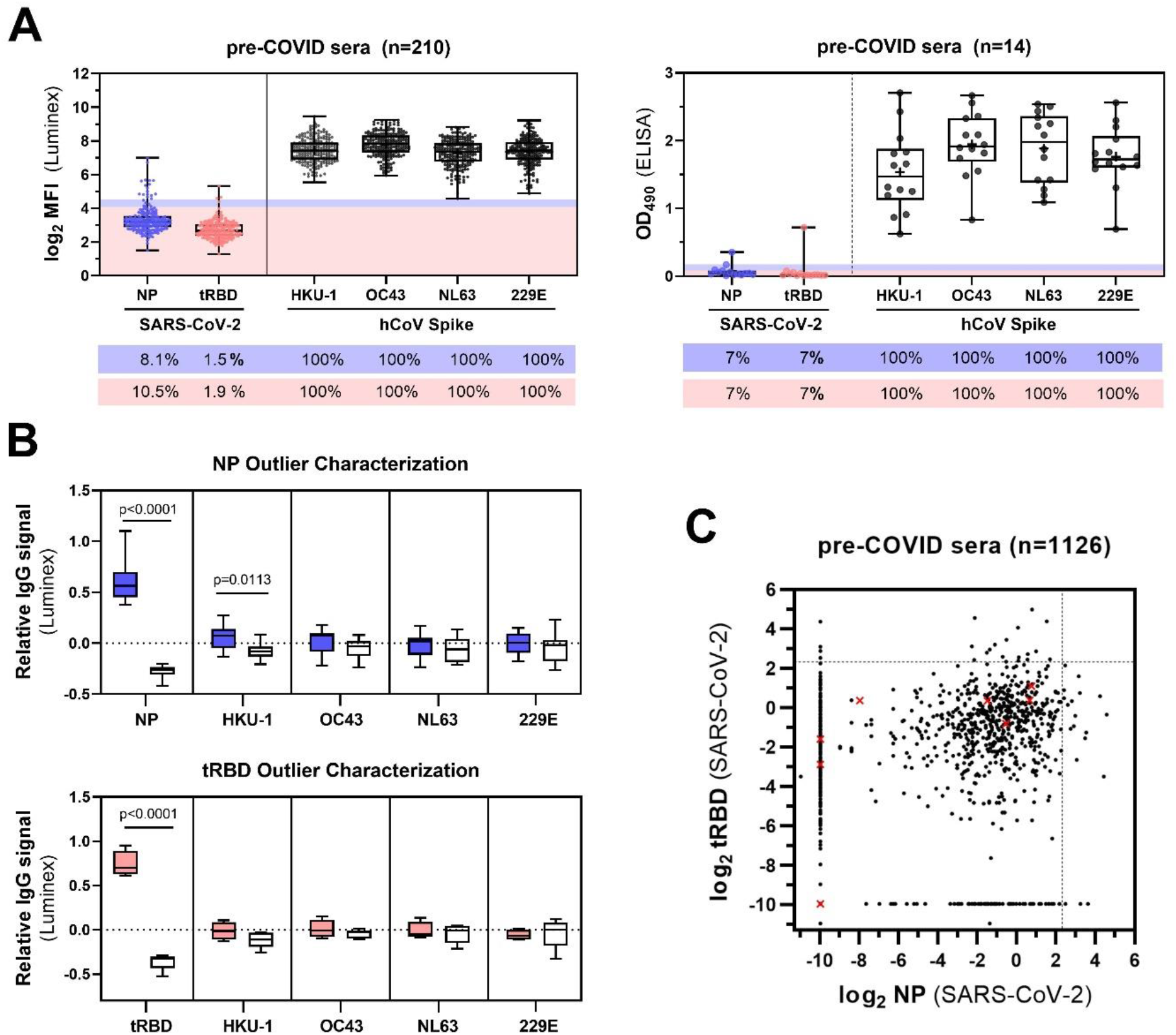
Characterization of cross-reactive IgG responses between SARS-CoV-2 and endemic hCoV strains in the employed specificity cohorts. **A)** Seroreactivity of serum samples from the two specificity cohorts (AIT pre-COVID-19 cohort, n=210 and MedUni Wien Biobank pre-COVID-19, n=14) employed for pre-validation of the SARS-CoV-2 tRBD and NP antigens with the Luminex or ELISA assays respectively, was measured with the spike proteins of common-cold hCoVs HKU-1, OC43, 229E and NL63. Outliers were classified as observations that fall above the 75^th^ percentile + 1.5 x IQR. Shades give the respective calculated cut-offs and are color-coded for NP (blue) or tRBD (pink). Values below the box-plots give the measured seroreactivity above these cut-offs in percent. **B)** Relative IgG levels of NP (n=17, blue boxes) and tRBD (n=4, pink boxes) outliers towards the spike proteins of hCoV. White boxes give relative IgG levels of sera with readouts <25^th^ percentile (n=16 for NP, n=5 for tRBD) to compare with outliers. Means within groups were compared by One-Way ANOVA followed by a Sidak test to correct for multiple comparisons. **C)** tRBD and NP-specific seroreactivity of the specificity cohort (n=1,126 MedUni Wien Biobank) used for clinical validation. Red crosses display sera from individuals with PCR-confirmed hCoV infection. Dashed lines indicate the cut-off of 5 U/mL.

### Clinical evaluation of test performance after symptom onset

Diagnostic accuracy of the Technozym NP or RBD IgG Tests was evaluated at different time points after symptom onset in plasma from hospitalized individuals (general ward and intensive care unit [ICU] patients) and outpatients. A total of 104 plasma samples were drawn during the acute and early convalescent phase of SARS-CoV-2 infection. NP-specific IgG levels correlated well with tRBD-specific IgG levels, even at levels being below the set threshold for seropositivity (1-5 d: r_s_=0.67, p<0.0001; 6-10 d: 0.76, <0.0001; 11-15 d: 0.76, 0.0006, **Fig. 5**). The positivity rates increased over time, peaking at 100% 15-22 days after symptom onset in both assays. True positivity rates for the NP ELISA were consistently higher than with the tRBD ELISA at all time points (1-5 d: NP vs tRBD: 14.7% vs 5.9%; 6-10 d: 45.7% vs 34.2%; 11-15 d: 76.5% vs 64.7%, **Fig. 5 and Table S2)**. Yet, sera displayed a great heterogeneity in antibody levels throughout the observation period (**Table S2**). None of the false-negative results among the samples were obtained with both assays. Astonishingly, 85.7% of the sera already contained neutralizing antibodies (median titer: 1:24; range 1:4 – 1:128, **Table S2**) as soon as by day 5 after symptom onset. Of these, however, only a total of 18% of the sera demonstrated seroreactivity above the cut-off for either the NP or tRBD antigen (**Fig. 5**). Yet, the quantitative nature of the assay allowed us to correlate antibody levels below the cut-off for seropositivity and we could demonstrate excellent correlation of tRBD-specific antibodies with neutralizing function at all four investigated time points (1-5 d: r_s_=0.49, p=0.0004; 6-10 d: 0.77, <0.0001; 11-15 d: 0.82, <0.0001; 16-22: 0.67, 0.0003, **Fig. 5**).

**Fig. 5.**
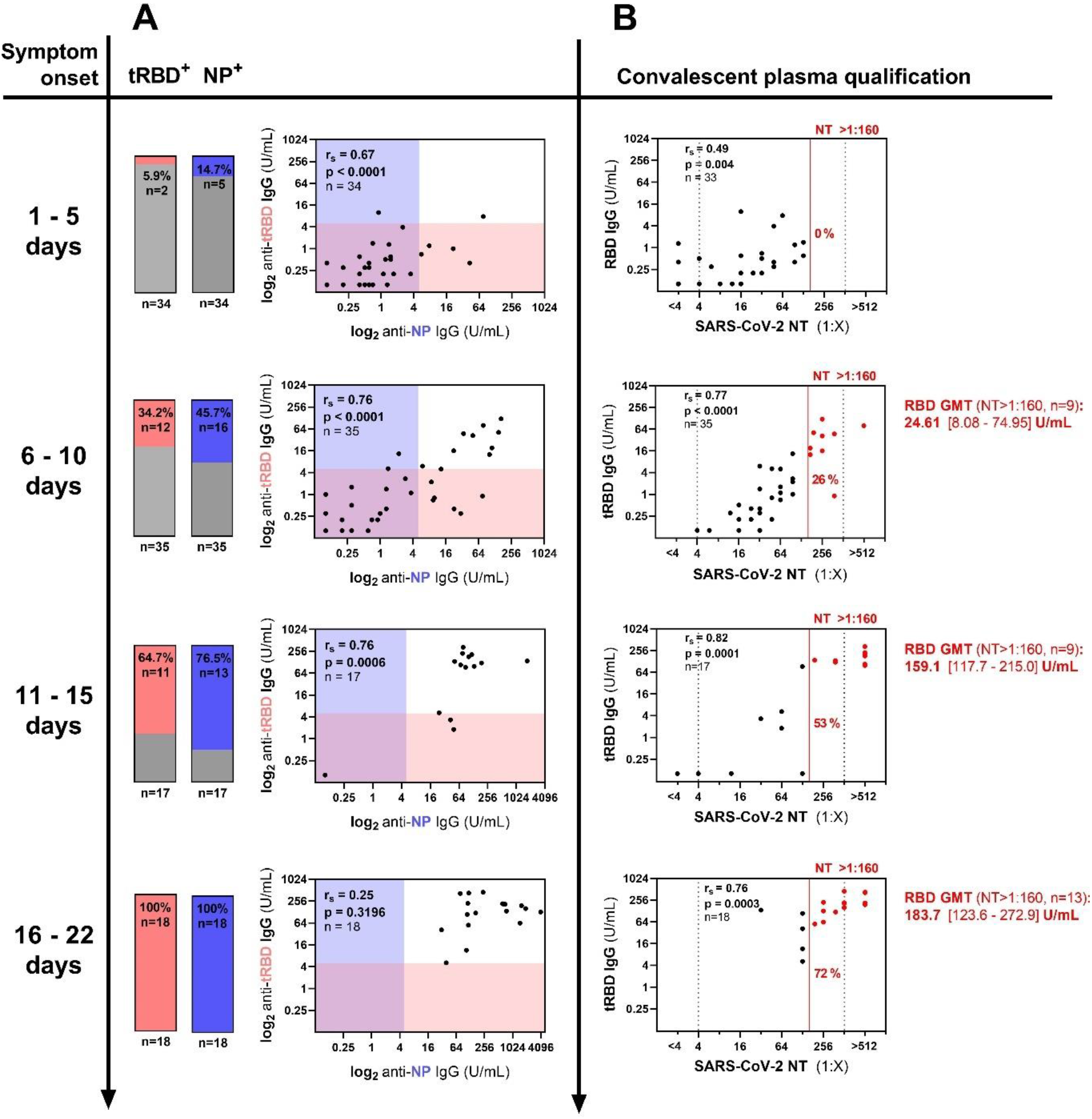
Time-resolved evaluation of NP, tRBD-specific and neutralizing antibodies in the acute and early convalescent phase after SARS-CoV-2 infection. **A-B**, A total of 104 plasma samples from 64 outpatients (16%) and hospitalized individuals (65% general ward, ICU 19%) were analyzed for anti-NP and anti-tRBD antibodies and neutralizing antibodies at the indicated time points. **A)** Antibody levels were assessed with the Technozym ELISAs according to the suggested cut-off of 5.000 U/mL. Bars indicate the fraction of NP, tRBD-positive samples among the tested. Shades give the respective ELISA cut-offs. **B)** Neutralization assays with authentic SARS-CoV-2 virus were performed within a serum dilution range of 1:4 – 1:512 (dashed lines). Values below or above these limits were assigned to 1:2 or 1:1,024 for correlation analysis, respectively. The red line indicates a NT of 1:160 that is recommended by the FDA for the screening of recovered COVID-19 patients for convalescent plasma therapy. All sera above this cut-off are color-coded in red. Geometric mean titers and 95% CI in the RBD ELISA are given for sera with a NT >1:160. r_s_, Spearman’s correlation factor.

## 3. Discussion

Superb assay specificity is of utmost importance for the assessment of antibodies directed against SARS-CoV-2, as a substantial proportion of infected individuals escapes identification due to the frequent asymptomatic course of the disease, thereby distorting the true humoral seroprevalence in any given population (*11*). The biological basis for false-positives is multifactorial, but the influence of the production platform and process-related peculiarities or impurities on protein performance are factors that are often underestimated. While the viral NP is almost always produced in bacteria (*12, 13*), we expressed the spike receptor binding domain in HEK cells, CHO cells, insect cells and plants (*4, 14*–*16*). To find out which of these systems leads to the highest quality and manufacturability of the RBD diagnostic antigen of potentially high demand, we evaluated these production platforms and pre-validated the proteins based on diagnostic performance with a large set of pre-COVID-19 and COVID-19 sera using the Luminex platform. All five expression platforms demonstrated suitability for the production of functional protein, proven by a binding assay with the SARS-CoV-2-RBD-specific mAb CR3022. Yet, in part due to the different transfection methods used, RBD yields from CHO-K1, CHO-S as well as from *Tnms*42 insect cells and tobacco plants were insufficient for sustainable commercial antigen production (< 1mg/L, **Fig. 1**). In contrast, HEK cells readily produced overall yields of 40 mg/L using PEI-transfection. Yields of 30 mg/mL per liter have also been described for CHO-expressed RBD. However, this can be traced back to optimized design of expression constructs and improved production processes for stable RBD-expressing CHO cells together with less extensive purification protocols (*17*). We observed higher basal seroreactivity of control sera with insect-derived RBD than with RBD from human and non-human mammalian cell lines; which is in line with other reports (*4*). Host-related impurities do not account for that, as insect-cell produced RBD demonstrated the highest purity among all our RBD samples (99%, **Fig. S2**). While there was a common set of false-positive samples shared by RBD from non-human and human mammalian cell lines as well as plants, false-positives reactive with the insect material were entirely insect-RBD-specific (**Fig. S6**). A possible reason may be platform-specific protein modifications, such as glycosylation, that provide the protein with a unique process-derived signature. Indeed, *T. ni*-derived insect cells were demonstrated to generate core α1,3-fucose structures with allergenic potential in humans (*18*), which might be associated with this peculiar seroreactivity profile.

Based on our observation that RBD tends to form homodimers in an unpredictable manner among different production batches of the same expression host, we used an optimized, truncated version of an RBD as diagnostic antigen -tRBD-, enabling the production of large amounts of RBD with consistent quality (**Fig. S2**). For tRBD performance, antigen purity was of utmost importance, even when expressed in human cell lines. At a consensus specificity of 99.1%, a reduction in tRBD purity by 10% (pure: 97.5%, impure: 87.5% purity) resulted in a drastic reduction in sensitivity by 83.9% (pure: 95.2% versus impure: 11.3%, respectively) in the Luminex pre-validation assays (**Fig. S4**). Since purity after an IMAC capture step was highly batch-dependent and resulted in inconsistent seroreactivity profiles, our standard downstream process included a scalable AIEX chromatography polishing step to account for these inconsistencies and to improve the diagnostic performance of the antigens.

The two test antigens, tRBD from HEK cells and NP from *E. coli*, were further used for ELISA assay development. We configured the assays with a number of sera taken from SARS-CoV-2-infected individuals with weak antibody responses to ensure high assay sensitivity. In contrast to available literature, we used high antigen coating concentrations (6 µg/mL) to yield satisfactory readouts (*4, 19, 20*) and to achieve a high dynamic measurement range. A caveat of many assay validation studies is that performance characteristics are skewed by the exclusive inclusion of samples from hospitalized individuals, where robust antibody levels are to be expected (*21*). Likewise, the sole consideration of healthy donors in control groups may lead to overestimated assay specificity, as the impact of potential cross-reactive factors present in the general population is largely ignored. In this respect, auto-antibodies commonly found in individuals with inflammatory diseases (*22*) were already described to cross-react with SARS-CoV-1 antigens (*23*). To challenge our tests systems, we biased our large specificity cohort (n=1,126) by including samples with an increased propensity for cross-reactivity, including sera from individuals with inflammatory illnesses (n=359), sera from PCR-confirmed hCoV infections (n=8) and sera drawn during winter months to increase the likelihood of respiratory infections (n=494). Similarly, our sensitivity cohort (n=244) included convalescent sera from SARS-CoV-2-infected individuals covering the full spectrum of clinical manifestations (from asymptomatic to ICU patients). Among them 21% of the sera were collected from asymptomatic individuals or from individuals with mild to moderate illness, who may mount less robust and durable antibody responses after an infection (*24*). Based on these cohorts, we defined adequate test parameters to enable highly specific detection of SARS-CoV-2-specific antibodies. A cutoff deduced by the 99^th^ percentile method (8.000 U/mL) allowed for high specific serodiagnosis with 99.2% for the Technozym RBD Test and 99.1% for the Technozym NP Test (at sensitivities of 86.3% and 76.7%, respectively). This is a remarkable result for a tetramethylbenzidine-based manual test system, considering the highly diverse nature of our study cohorts. While some automated systems were described to achieve specificities approximating 100% (*25, 26*), assay performance is highly cohort-specific. The use of diverse study cohorts was also associated with performance deteriorations in such test platforms (i.e. Abbot, Specificity: 97.5%)(*27*). For the Meduni Wien Biobank cohort we had performance data with CE-marked automated test systems available (*10*) to directly compare with our ELISAs at the high specificity cut-off criterion (8.000 U/mL). With an AUC of 0.987 [0.979-0.992] and a specificity of 99.1%, the NP ELISA presented with comparable performance to the Abbott SARS-CoV-2 chemiluminescence microparticle assay (AUC: 0.993 [0.987-0.997], **Fig. S5**, Sp 99.2%) (*10*), that also relies on the NP antigen. The tRBD ELISA even outperformed the DiaSorin LIAISON^®^ SARS-CoV-2 S1/S2 IgG chemiluminescence assay (tRBD ELISA: AUC/Sp/Sen=0.993/99.2%/84.9% vs DiaSorin:0.976/98.2%/82.8%, see **Fig. S5** and Perkmann *et al* (*10*). While we cannot rule out minor cross-reactivity between hCoV-specific antibodies and SARS-CoV-2 antigens, they appeared to have a limited effect on assay performances (**Fig. 4C**).

Yet, for an estimated seroprevalence of 5% in the general European population (*28, 29*), a test with a specificity and sensitivity of 99.2% and 86.3%, respectively, only scores a PPV of 85.0% resulting in 15 false-positive results out of 100, which is still insufficient. In line with previous results from us and others (*30*–*32*), we demonstrate that false-positive results are largely antigen-dependent (**Fig. 2B, Fig. 4C**). Orthogonal testing is suggested by the Centers of Disease Control and Prevention (CDC) to remedy specificity problems in low transmission settings (*33*). Previous studies have used RBD as screening antigen and the trimeric spike protein or the spike S2 domain in second-line tests to confirm initial positive results (*4, 32*). Such conventional orthogonal test strategies, however, increase specificity often at the expense of sensitivity. We therefore established an adaptive orthogonal test algorithm where positive sera were first identified with the tRBD ELISA allowing for highly sensitive testing (at the expense of specificity) and samples within a predefined area of uncertainty then underwent confirmatory testing with the NP ELISA (*10*). This two-test algorithm resulted in a cumulative specificity of 99.8% and an even higher sensitivity of 88.1% (+0.037, p<0.050),yielded a PPV of 96.3% [86.7-99.1] (**Fig. 3**). This is an excellent result for a manual test format and its specificity is on par with other orthogonal tests relying on automated systems (*10*).

As the Technozym NP and RBD ELISAs provide a five-point calibrator, set ELISA antibody levels can be quantified, compared and followed over time. For such an application, we chose a cut-off of 5.000 U/mL that allowed for more sensitive analysis of antibody levels at acceptable specificity, adapted from the cut-off given by the Youden index. With convalescent sera taken at median 43-54 days post-symptom onset, the tRBD ELISA allowed for a more sensitive detection of antibodies than the NP ELISA (**Fig. 3A, B**). Yet, time-resolved analysis of seroconversion demonstrated that NP-specific antibodies develop earlier after an infection and true positive rates were consistently higher with the NP ELISA for samples collected within the first 15 days post-symptom onset (**Fig. 5, Table S2**). This phenomenon has already been described in patients infected with SARS-CoV-1(*34, 35*) and was associated with higher sensitivities of other SARS-CoV-2 test systems, relying on the NP, in the early phase after an infection (*36*). Determining the neutralizing capacity of SARS-CoV-2 anti-RBD antibodies is critical to elucidate possible protective effects of the immune response. Considering all neutralizing activity above background as positive, we observed neutralizing antibodies in 85% of the sera already by day five after symptom onset (**Fig. 5**), which is in line with previous studies (*37, 38*). Of note, RBD-seroconversion, defined by antibody levels above a threshold of 5.000 U/mL, was observed for only 6% of the sera at this time point. Yet, despite 33 out of 35 samples demonstrating reactivity below our pre-defined cutoff, neutralizing titers correlated well with RBD-specific IgG responses. A recent study demonstrated that the early neutralizing response is dominated by RBD-specific IgA antibodies (*39*). As we exclusively measured RBD-specific IgG responses we cannot rule out that part of the early neutralizing activity we observe derive from neutralizing IgA or even earlier IgM responses.

Tests for the screening of reconvalescent COVID-19 patients for the presence of anti-SARS-CoV-2 antibodies are of great interest for identifying suitable donors for convalescent plasma therapy (*40*). A retrospective, propensity score–matched case–control study performed at the Mount Sinai hospital (New York, NY) provides evidence for a survival benefit in patients receiving convalescent plasma transfusion as an effective intervention in COVID-19 (*40*). In August 2020, the FDA issued a new guidance on the Emergency Use Authorization (EUA) for COVID-19 convalescent plasma, recommending plasma donations to be qualified by either the Mount Sinai COVID-19 ELISA IgG Antibody Test or Ortho VITROS IgG assay (*41*). Prior to this guidance, NTs of at least 1:160 were considered acceptable in the absence of high-titer samples (*42*). As we did not have the beforementioned tests available, we qualified plasma donors according to the NT 1:160 criterion. The fraction of samples exceeding this threshold gradually increased over time and by day 15 after symptom onset, 53% of the sera and by day 22, 72% of sera had titers higher than 1:160 (**Fig. 5, Table S2**). The geometric mean RBD titers in these sera corresponded to 159.1 U/mL and 183.7 U/mL, respectively. Since correlates of protection from infection remain to be determined we cannot deduce whether these titers are clinically relevant in prophylaxis, at this point.

## 4. Discussion

In conclusion, we have developed two highly specific, quantitative, easy-to-implement and now commercially available SARS-CoV-2 antibody tests and defined optimal thresholds for their application in different aspects of clinical use. In addition to their simple format, the two tests are equally well suited as most automated CE-marked systems for high specificity applications, such as seroprevalence studies. Moreover, the RBD ELISA allows for the identification of donors for convalescent plasma therapy as RBD-specific antibody levels correlate well with the induction of functional neutralization responses. Both tests allow to comprehensively monitor the dynamics of antibody responses after infection. Yet, our data disclose different kinetics for antigen-specific antibody responses, which affect their performance at different time points after an infection. These findings are essential for ongoing efforts to establish serological tests for clinical diagnostics. In this respect, also test performance with convalescent sera collected more than 2 months after infection and the effect of antigen-specific antibody waning should be carefully addressed in future studies and compared to the comprehensive findings of this study.

## 5 Methods

### 5.1 Production of recombinant SARS-CoV-2 antigens for serodiagnosis

#### 5.1.1. Genetic constructs

pCAGGS mammalian expression vectors encoding the canonical SARS-CoV-2 receptor-binding domain (RBD, pCAGGS-RBD, aa Arg319 – Phe541, residue numbering as in NCBI Reference sequence: YP_009724390.1) sequence from the first human isolate Wuhan-1 (*43*) with a C-terminal hexa-histidine tag, were a kind gift from Florian Krammer, Icahn School of Medicine at Mount Sinai, NY (*4*). Both sequences were codon-optimized for the expression in mammalian cells.

A pTT28 mammalian expression vector (National Research Council, NRC, Ottawa, Canada) encoding a truncated version of the SARS-CoV-2 Spike receptor-binding domain (tRBD, pTT28-tRBD, aa Arg319 - Lys537) with a C-terminal octa-histidine tag was generated.

A pEAQ-HT plant expression vector (*44*) encoding RBD (pEAQ-HT-RBD, aa Arg319 – Phe541) fused to the barley α-amylase signal peptide and a C-terminal hexa-histidine tag was generated. The RBD sequence was codon-optimized for the expression in plants and synthesized by GeneArt (Thermo Fisher Scientific, Regensburg, DE).

A pET30a*cer E. coli* expression vector (*8*) encoding the full-length SARS-CoV-2 Wuhan-1 NP sequence (aa Met1–Ala419, GenBank: NC_045512.2) (*43*) fused to a completely removable N-terminal CASPON tag (*7, 45*), yielding pET30a*cer*-CASPON-NP, was generated as described elsewhere (*9*). Briefly, SARS-CoV-2 NP sequence was amplified via PCR using the qPCR positive control plasmid 2019-nCoV_N obtained from Integrated DNA Technologies (Coralville, Iowa, USA) and was fused to the CASPON tag consisting of the negative charged T7AC solubility tag (*7*), a hexa-histidine tag, a short linker (GSG) and the caspase-2 cleavage site (VDVAD) resulting in the sequence MLEDPERNKERKEAELQAQTAEQHHHHHHGSGVDVAD.

Expression vectors pFUSEss-CHIg-hG1 and pFUSEss2-CLIg-hK, encoding the heavy and light chains of the SARS-CoV/SARS-CoV-2 monoclonal antibody CR3022 (*46*) were kindly provided by Florian Krammer (Icahn School of Medicine at Mount Sinai, New York, NY).

#### 5.1.2 Large-scale production of transfection-grade plasmid DNA

Plasmid DNA for transient transfection of HEK293-6E cells was produced according to an upstream process described previously (*47*). Briefly, the plasmids pCAGGS-RBD and pTT28-tRBD were transformed into *E. coli* JM108 by electroporation and cultivated in 1-L fed-batch mode. Cells were harvested by centrifugation and pDNA was extracted by alkaline lysis at 5 g/L cellular dry mass (CDM) following a protocol of Urthaler and colleagues (*48*). pDNA was processed to >95% purity by multiple chromatography steps based on a platform purification protocol (Cytiva, Little Chalfont, UK)(*49*).

#### 5.1.3 Transient expression of RBD, tRBD and NP in diverse biotechnological platforms

##### Human embryonic kidney cells

###### Shake flask cultivation

HEK293-6E cells (licensed from National Research Council, NRC, Ottawa, Canada) were routinely cultivated in suspension in Freestyle™ F17 medium supplemented with 4 mM L-glutamine, 0.1% (v/v) Pluronic F-68 and 25 µg/mL G-418 (all Thermo Fisher Scientific, Waltham, MA) in a humidified atmosphere of 5-8% (v/v) CO_2_ at 37°C shaking at 125 rpm. Polyethylenimine (PEI)-mediated transient transfections with either pCAGGS-RBD, pTT28-tRBD or pFUSEss-CHIg-hG1 and pFUSEss2-CLIg-hK for the expression or RBD, tRBD or mAb CR3022 were performed according to the manufacturer’s protocol as previously described (*50, 51*).

Transfections were performed by dropwise addition of a mixture of one µg plasmid DNA and two µg linear 25-kDa or 40-kDa PEI (Polysciences, Inc., Hirschberg, DE) per mL of culture volume (1.7 – 2.0 x 10^6^ cells/mL). Two and four days post-transfection, cells were supplemented with 0.5% (w/V) tryptone N1 (Organotechnie, La Courneuve, FR) and 0.25% (w/V) D(+)-glucose (Carl Roth, Karlsruhe, DE). Supernatants were harvested five to six days post-transfection by centrifugation (2000 g, 15 min) and were filtered through 0.45 µm filters before downstream procedures.

###### Medium-scale cultivation

Stepwise upscaling was performed using a Multi-bioreactor system DASGIP (Eppendorf, Hamburg, DE) followed by a 10 liter scale bioreactor System BioFlo320 (Eppendorf, Hamburg, DE). The bioreactors were inoculated at half the final volume (F17 expression medium supplemented with 4 mM L-Glutamine and 0.1% Pluronic) with a seeding density of 0.5 x 10^6^. cells/mL. The inoculum was prepared in shake flask cultures as described above. The bioreactors were controlled to a pH of 7.2 using CO2 and 7.5% (w/V) carbonate base and to 50% (v/v) dissolved oxygen by submerged aeration. Transfection was performed at a cell concentration of 1.7 x 10^6^. PEI and the respective plasmid DNA were diluted in media mixed and incubated at room temperature for 10 minutes prior to addition to the cultures (45 µg PEI and 15 µg of plasmid per 10^6^ cells). Twenty four hours post transfection, cells were expanded and 24 hours later were fed a TN1 peptone at a concentration of 0.5% (v/v). Each day post-transfection viability, cell density and glucose concentration were measured and a daily bolus feed to a glucose concentration of 2.5 g/L was performed. The cultures were harvested once viability dropped below 60%.

##### Chinese hamster ovary cells

CHO-K1 and CHO-S cells were routinely propagated in CD-CHO medium (Thermo Fisher Scientific, Waltham, MA) or in Hyclone Actipro medium (Cytiva, Chicago, IL) both supplemented with 0.2% (v/v) Anti-Clumping Agent (Thermo Fisher Scientific, Waltham, MA) and 8 mM L-glutamine (CHO-K1, Sigma Aldrich, St. Louis, MO) or 8 mM GlutaMAX (CHO-S, Thermo Fisher Scientific, Waltham, MA), respectively. Cells were cultivated in suspension at 37°C, 7% (v/v) CO_2_ and humidified air, shaking at 140 rpm.

For nucleofection, a total of 1 x 10^7^ cells in the exponential growth phase were pelleted for 8 min at 170 g and were resuspended in 99 μL resuspension buffer R (Thermo Fisher Scientific, Waltham, MA). Cells were mixed with pCAGGS-RBD, which had been pre-diluted with UltraPure™ DNase/RNase-Free distilled water to a concentration of 2 µg/µL in a total volume of 11 µL and were electroporated with a Neon^®^ Nucleofector using a 100 μL Neon^®^ Transfection Kit (all Thermo Fisher Scientific, Waltham, MA) with 1700 V and one pulse of 20 ms. Seven to eight transfections per cell line were performed and subsequently pooled in a 500 mL shake flask with a 200 mL working volume. Supernatants were harvested five days post transfection by centrifugation (170 g, 10 min) and were sterile-filtered before further use.

##### Insect cells

*Tnms*42, an alphanodavirus-free subclone of the High-Five insect cell line (*52, 53*), were routinely propagated in adherent culture in HyClone SFM4 insect cell medium (Cytiva, Marlborough, MA) at 27°C and were expanded in suspension culture for recombinant protein expression. A passage one virus seed stock expressing the SARS CoV-2 RBD was amplified in *Sf*9 cells to generate a passage three working stock and was titrated by plaque assay as previously described (*54*). *Tnms*42 insect cells at 2 x 10^6^ cells/mL were infected at a multiplicity of infection (MOI) of two, and the supernatant was harvested three days post-infection, clarified (1,000 g, 10 min, followed by 10,000 g, 30 min) and was filtered through a 0.45 µm filter before downstream procedures.

##### Tobacco plants

The pEAQ-HT-RBD expression vector was transformed into *Agrobacterium tumefaciens* strain UIA143 (*44*). Syringe-mediated agroinfiltration of leaves from five-week-old *Nicotiana benthamiana* ΔXT/FT plants was used for transient expression (*55*). Four days after infiltration, leaves were harvested and intracellular fluid was collected by low-speed centrifugation as described in detail elsewhere (*56*).

##### E. coli

The pET30a*cer*-CASPON-NP expression vector was transformed into *E. coli* enGenes-X-press for growth-decoupled recombinant protein production as described elsewhere(*8*) Briefly, for cultivation cells were grown in fed-batch mode in a 1.0 L (0.5 L batch volume, 0.5 L feed) DASGIP^®^ Parallel Bioreactor System (Eppendorf AG, Hamburg, DE) equipped with standard probes (pH, dissolved oxygen [pDO]). The pH was maintained at 7.0 ± 0.05, temperature was maintained at 37 ± 0.5°C during the batch phase and decreased to 30 ± 0.5°C at the beginning of the feed phase. The dissolved oxygen level was stabilized at > 30% (V/V). Induction of NP production was facilitated at feed hour 19 with the addition of 0.1 mM IPTG and 100 mM arabinose.

#### 5.1.4. Downstream procedures

##### Purification of SARS-CoV-2 RBD and tRBD from different expression systems

His-tagged RBD and tRBD from filtered HEK supernatants, as well as RBD from *Tnms*42 insect cell supernatants, were concentrated and diafiltrated against 20 mM sodium phosphate buffer supplemented with 500 mM NaCl and 20 mM imidazole (pH 7.4) using a Labscale TFF system equipped with a Pellicon™ XL Ultracel 5 kDa, 0.005 m^2^ ultrafiltration module (Merck, Darmstadt, DE). The proteins were captured using a 5-mL HisTrap FF Crude or a 1-mL HisTrap Excel immobilized metal affinity chromatography (IMAC) column connected to an ÄKTA Pure chromatography system (all from Cytiva, Marlborough, MA) and were eluted by applying a linear gradient of 20 to 500 mM imidazole over 5 to 20 column volumes, as appropriate. Intracellular fluid collected from plant material was directly loaded onto a 5-mL HisTrap HP column and was purified as described elsewhere (*57*). CHO-K1 and CHO-S expression supernatants were supplemented with 20 mM imidazole and were directly loaded onto a 1-mL HisTrap FF column connected to an ÄKTA Start chromatography system (both Cytiva, Marlborough, MA), equilibrated with 50 mM sodium phosphate buffer supplemented with 300 mM NaCl and 20 mM imidazole (pH 7.4). Proteins were eluted by applying a linear gradient of 20 to 500 mM imidazole over 20 column volumes.

Fractions containing RBD or tRBD were pooled and either diluted with 20 mM sodium phosphate buffer (pH 7.4) to a conductivity of ∼10 mS/cm and then loaded onto a Fractogel EMD DEAE column (Merck Millipore, Germany) or loaded onto HiTrap DEAE FF column (Cytiva, Marlborough, MA), both pre-equilibrated with 20 mM sodium phosphate buffer _(_pH 7.4). A residence time of 2 minutes was used. The flow-through fractions, containing RBD or tRBD, were collected. Impurities were subsequently eluted using 20 mM sodium phosphate buffer, 1 M NaCl, pH 7.4 and the column was cleaned in place by incubation in 0.5 M NaOH for 30 minutes. The protein of interest present in the flow-through fraction was buffer-exchanged into PBS using Amicon Ultra-15 Ultracel 10 kDa spin columns (Merck Millipore, Germany) or was dialyzed against PBS. IMAC-captured RBD from insect cell supernatants was ultra- and diafiltrated using Amicon Ultra Centrifugal Filter Units (10 kDa MWCO, Merck Millipore) to change the buffer to PBS and was further purified by size exclusion chromatography using a HiLoad 16/600 Superdex 200 pg column (Cytiva, Marlborough, MA) equilibrated with the same buffer. Fractions containing RBD were concentrated using Amicon Ultra-15 Ultracel 10 kDa spin columns (Merck Millipore, Germany). All purified proteins were quantified by measuring their absorbance at A_280_ with a Nanodrop instrument and stored at −80°C until further use.

##### Purification of SARS-CoV-2 NP from *E. coli* cellular lysates

The purification of NP was optimized and performed as described by De Vos and colleagues (*9*). In brief, NP was produced by using the CASPON platform process(*45*) with modifications. The process consisted of an IMAC capture step (WorkBeads 40 Ni NTA, Bio-Works, Uppsala, SE) of the clarified cell lysate. A nuclease treatment (Salt Active Nuclease High Quality, ArcticZymes Technologies ASA, Tromsø, NO) was required to reduce CASPON-NP nucleic acid binding. Imidazole was removed from the IMAC eluate using a Butyl Sepharose HP hydrophobic interaction chromatography (Cytiva, Uppsala, SE) which also separated full-length from fragmented CASPON-NP. A variant of cpCasp2 (*7*) was used to remove the affinity fusion-tag. Finally, an IMAC polishing step was used to separate native NP from residual CASPON-NP, the free affinity fusion-tag, the affinity-tagged cpCasp2 variant and metal binding host cell proteins. The polishing fraction was buffer exchanged to PBS using tangential flow filtration on Pellicon 3 Ultracel 10 kDa membrane (Merck Millipore, Darmstadt, DE).

##### Purification of mAb CR3022

mAb CR3022 was purified by affinity chromatography using a 5-mL HiTrap Protein A HP column connected to an ÄKTA pure chromatography system (both from Cytiva, Marlborough, MA) according to the manufacturer’s protocol. The antibody was eluted using 0.1 M glycine-HCl buffer (pH 3.5). Eluate fractions containing CR3022 were immediately neutralized using 1 M Tris-HCl buffer (pH 8.0), pooled and concentrated using Amicon ultrafiltration cartridges with a cut-off of 10 kDa (Merck, Darmstadt, DE) and were further dialyzed against PBS (pH 7.4) at 4°C overnight using SnakeSkin Dialysis Tubing with a 10 kDa cut-off (Thermo Fisher Scientific, Germering, DE). CR3022 was further purified by size exclusion chromatography using a HiLoad 16/600 Superdex 200 pg column (Cytiva, Marlborough, MA) equilibrated with the same buffer as used for dialysis.

### 5.2 Commercial antigen and antibody reagents

Recombinant spike proteins of the four common cold hCoV strains, HKU-1, OC43, NL63 and 229E were purchased from Sino Biological Inc, Beijing, CN (#40606-V08B, #40607-V08B, #40604-V08B and #40605-V08B, respectively). A recombinant chimeric human/mouse anti-SARS-CoV-2 NP antibody consisting of a mouse scFv fused to the Fc region of human IgG1 (clone 1A6) was purchased from Abcam, Cambridge, UK (#ab272852).

### 5.3 Assessment of recombinant protein quality

#### 5.3.1 Analytical size exclusion chromatography (SEC)

High-performance liquid chromatography (HP)-SEC experiments were performed on a Dionex™ UltiMate™ 3000 RSLC system equipped with an LPG-3400SD Standard Quaternary Pump module, a WPS-3000 TSL Analytical Split-Loop Well Plate Autosampler and a DAD-3000 Diode Array Detector equipped with a ten µL analytical flow cell (all from Thermo Fisher Scientific, Germering, DE). RBD, tRBD and NP samples (25-80 µg per sample) were run on a Superdex™ 200 Increase 10/300 GL column (Cytiva, Uppsala, SE) and UV signals were detected at λ = 280 nm. For RBD and tRBD, Dulbecco’s PBS buffer (DPBS) supplemented with 200 mM NaCl was used as mobile phase, the flow rate was set to 0.75 mL/min and a 45 min isocratic elution was performed. For NP samples 0.1 M sodium phosphate buffer (pH 7.0) containing 300 mM NaCl was used as mobile phase, the flow rate was set to 0.5 mL/min and a 60 min isocratic elution was performed. HP control, data acquisition and data evaluation were performed using Chromeleon™ 7.2 Chromatography Data System software (Thermo Fisher Scientific, Germering, DE). Sample purity (P), monomer (M), dimer (D) and full-length (FL) content were determined based on the respective peak area of the UV signal at 280 nm. For RBD and tRBD purity was defined as P=(M+D)/total area, monomer and dimer content were respectively defined as M[%]=M/(M+D)∗100 and D=100-M[%]. For NP, full-length content was defined as FL[%]=FL/total area.

#### 5.3.2 Bio-Layer Interferometry (BLI) measurements

Interaction studies of RBD, tRBD and NP with in-house produced anti-RBD mAb CR3022 and a commercial anti-SARS-CoV-2 nucleocapsid protein antibody (ab272852, Abcam, Cambridge, UK) were performed on an Octet RED96e system using high precision streptavidin (SAX) biosensors (both from FortéBio, Fremont, CA). Antibodies were biotinylated using the EZ-Link Sulfo-NHS-LC-Biotin kit (Thermo Fisher Scientific, Waltham, MA). Excess sulfo-NHS-LC-biotin was quenched by adding Tris-HCl buffer (800 mM, pH 7.4) to a final concentration of 3 mM. Biotinylated antibodies were further purified using PD-10 desalting columns (Cytiva, Marlborough, MA) according to the manufacturer’s protocol. All binding assays were conducted in PBS supplemented with 0.05% (v/v) Tween 20 and 0.1% (w/V) BSA (PBST-BSA) at 25°C with the plate shaking at 1,000 rpm. SAX biosensors were first equilibrated in PBST-BSA and then loaded with the respective biotinylated capture molecules, either for 180 sec (34 nM CR3022 solution) or until a signal threshold of 0.8 nm was reached (50 nM anti-NP mAb solution). Subsequently, antibody-loaded biosensors were dipped into PBST-BSA for 90 sec to record a baseline, before they were submerged into different concentrations of their respective analytes. To determine *K*_D_ values, biotinylated antibodies loaded onto biosensors were exposed to six concentrations of the binding partners (RBD, tRBD or NP) to cover a broad concentration range around the respective *K*_D_ value (*58*). For antigen association, mAb CR3022 was exposed to a three-fold serial dilution of RBD or tRBD (range: 300 nM - 1.2 nM in PBST-BSA) for 300 sec, while anti-NP mAb-was dipped into two-fold serial dilutions of the NP protein (40 nM - 1.3 nM in PBST-BSA) for 600 sec. For dissociation, the biosensors were dipped into PBST-BSA. Each experiment included a baseline measurement using PBST-BSA (negative control) as well as a positive control (RBD monomer) where applicable. SAX biosensors loaded with biotinylated CR3022 or anti-NP mAb could be regenerated by dipping them into 100 mM glycine buffer (pH 2.5). RBD or tRBD proteins were measured in triplicates or quadruplicates, while NP proteins were measured in duplicates. No unspecific binding of proteins to SAX biosensors was observed. Data were evaluated under consideration of the lower limit of detection (LLOD) and lower limit of quantification (LLOQ) as reported elsewhere (*59, 60*). The analysis was performed using the Octet data analysis software version 11.1.1.39 (FortéBio, Fremont, CA) according to the manufacturer’s guidelines. For easier comparison of the RBD variants produced in different expression hosts, the *K*_D_ values were determined from the measured equilibrium response (steady state analysis). However, the interaction between the CR3022 mAb and the final tRBD batches were also evaluated kinetically by fitting the BLI data to a 2:1 heterogeneous ligand binding model. Note, although the CR3022 mAb has two identical binding sites, the second binding event is dependent on the first binding since allosteric effects or sterical hindrance can ultimately lead to a positive or negative cooperative binding behavior (*51, 61, 62*). However, in case of the reported interaction, the affinity constant (*K*_D_) values are very close to one other in the low nanomolar range.

The interaction between the NP protein and the anti-NP mAb is difficult to characterize due to avidity effects that arise from the dimeric nature of both interaction partners. Kinetic evaluation of the BLI data is problematic since the dissociation curves are heterogenic. Additionally, if the dissociation phase shows less than 5% decrease in signal during the defined dissociation phase, as observed for the lower concentration range of NP protein, a precise determination of the dissociation rate constants (*k*_d_) is not possible (*63, 64*). However, it is feasible to calculate an upper limit for the *k*_d_ (s^-1^) which is given by *k*_d_<−ln(0.95)/*t*_d_, where td is the dissociation time in seconds (*63, 65*) Thus, an upper limit for the *K*_D_ value, calculated by the ratio of kd/ka, resulted in < 0.7 nM, suggesting a strong interaction in the picomolar range. Moreover, for comparison of single batches the observed binding rate (*k*_obs_) was plotted as a function of the NP concentration and used for the comparison of the single batches.

#### 5.3.3 Liquid Chromatography Electrospray Ionization Mass Spectrometry (LC/ESI-MS)

Purified proteins were S-alkylated with iodoacetamide and digested with endoproteinases LysC (Roche, Basel, CH) and GluC (Promega, Madison, WI) or chymotrypsin (Roche, Basel, CH) in solution. Digested samples were analyzed using a Thermo Ultimate 3000 HP connected to a 150 x 0.32 mm, 5 µm BioBasic C18 column (both Thermo Fisher Scientific, Waltham, MA) and a maXis 4G QTOF mass spectrometer (Bruker, Billerica, MA). An 80 mM ammonium format buffer was used as the aqueous solvent and a linear gradient from 5% B (B: 80% acetonitrile) to 40% B in 45 min at a flow rate of 6 µL/min was applied, followed by a 15 min gradient from 40% B to 95% B that facilitated elution of large peptides. The MS system was equipped with the standard ESI source and operated in positive ion, DDA mode (= switching to MSMS mode for eluting peaks). MS-scans were recorded (range: 150-2,200 Da) and the six highest peaks were selected for fragmentation. Instrument calibration was performed using ESI calibration mixture (Agilent, Santa Clara, CA). The analysis files were converted (using Data Analysis, Bruker) to mgf files, which are suitable for performing a MS/MS ion search with MASCOT. The files were searched against a database containing the target sequences. In addition, manual glycopeptide searches were done. Glycopeptides were identified as sets of peaks consisting of the peptide moiety and the attached N-glycan varying in the number of HexNAc, hexose, deoxyhexose and pentose residues. Theoretical masses of these peptides were determined using the monoisotopic masses for the respective amino acids and monosaccharides.

### 5.4 Ethics statement

The present study includes work with human sera from three different sites. Acute lithium heparin plasma samples collected from outpatient and hospitalized individuals for routine clinical testing were available at the B&S Central Laboratory Linz, Austria. Left-over samples were assessed for SARS-CoV-2 antibody levels and neutralizing titers in the early phase of infection and the study protocol was approved by the ethics committee of Upper Austria (EK1083/2020), in accordance with the Declaration of Helsinki. For ELISA validations, left-over sera from SARS-CoV-2 patients and sera from convalescent donors, as well as historical sera (<2020) were taken from the MedUni Wien Biobank, as approved by the ethics committee of the Medical University of Vienna (EK 1424/2020). The underlying sample collections were reviewed and approved by the ethics committee of the Medical University of Vienna (EK 595/2005, EK 404/2011, EK 518/2011), or by the ethics committee of the City of Vienna (EK-11-117-0711), respectively. Samples from hospitalized COVID-19 patients at the University Hospital of Innsbruck, reconvalescent COVID-19 patients with persistent cardio-pulmonary damage participating in a prospective observational study (CovILD-study, ClinicalTrials.gov number, NCT04416100, Reference: PMID: 33303539) and reconvalescent persons volunteering as plasma donors were used for test validation in Innsbruck (*66*). The underlying sample collections were reviewed and approved by the ethics committee of the Medical University of Innsbruck (EK 1103/2020, EK 1167/2020). Left-over SARS-CoV-2 acute and convalescent sera from blood donors and pre-COVID-19 sera from the Austrian Institute of Technology were taken for SARS-CoV-2 antigen pre-validation and the study was approved by the ethics committee of the city of Vienna (EK 20-179-0820).

### 5.5 Human serum and plasma samples

#### 5.5.1 Sensitivity cohorts

##### SARS-CoV-2 acute sera from a cohort of outpatient and hospitalized individuals, B&S Central Laboratory Linz, Austria

A cohort of hospitalized individuals and outpatients included a total number of 64 SARS-CoV-2 RT-PCR-confirmed (from respiratory specimens) COVID-19 patients (median age 65 [14-95, IQR 56-87 years], 17.2% females) who were treated in one of the two tertiary care hospitals Konventhospital Barmherzige Brueder Linz or Ordensklinikum Linz Barmherzige Schwestern in Linz, Austria, between March 15^th^ – April 10^th^ 2020. Of these, ten patients were treated as outpatients and 54 patients were hospitalized; twelve of them were treated at the intensive care unit (ICU). From the 64 patients, a total of 104 serial blood samples were drawn at different time points after symptom onset until April 10^th,^ 2020. Sixty-four patients had at least one, 28 patients had two, nine patients had three and three patients had four blood draws, which were sent to the central laboratory for routine clinical testing. The date of onset of symptoms was retrieved from medical records and was available for all patients. Left-over lithium heparin plasma samples were aliquoted and frozen at −80°C and had up to two freeze-thaw cycles.

##### Sera of SARS-CoV-2-positive patients and convalescent donors, Medical University of Vienna and Medical University of Innsbruck

The SARS-CoV-2 positive samples for ELISA validation comprise 70 serum specimens from unique patients or convalescent donors with (previous) SARS-CoV-2 infection from Vienna (either PCR-positive or symptomatic close contacts), as well as 174 SARS-CoV-2 PCR positive patients including hospitalized patients (n=123) and convalescent blood donors (n=51) from Innsbruck. All samples were collected >14 days after symptom onset (or positive PCR, in case of asymptomatic infection). A representative serum panel of these samples (n=28-31) was taken for the pre-validation of SARS-CoV-2 antigens by ELISA and for the assessment of SARS-CoV-2 neutralization titers.

##### SARS-CoV-2-convalescent and acute sera from a cohort of non-hospitalized blood donors, Austrian Institute of Technology (AIT) and Medical University of Vienna

The sensitivity cohort for antigen pre-validation covered 124 COVID sera. Among these, 96 sera were deidentified excess samples from infected individuals collected for routine SARS-CoV-2 serodiagnosis using a seven-plex bead-based Luminex-FlexMap system-based serotest and were available at the AIT. These serotests had been conducted similar to the analysis procedure outlined below. Seronegativity and/or seropositivity was based on cut-off values and end-point titers defined according to Frey *et al* (*67*) on the basis of 160 pre-COVID-19 sera. Additionally, the study cohort included a set of 28 COVID-19 sera from the Medical University (from the above), covering samples from primarily asymptomatic individuals or those with mild to moderate illness.

#### 5.5.2 Specificity cohorts

##### Pre-COVID-19 cohort, MedUni Wien Biobank

The pre-COVID-19 cohort covered a total of 1,126 samples from healthy, non-SARS-CoV-2-infected individuals collected before 2020 to guarantee seronegativity. Banked human samples including sera from voluntary donors (n=265, median age 38 [25-52] years, 59.0% females), samples from a large population-based cohort aged 8-80 years, representing a cross-section of the Austrian population (N=494, collected 2012-2016 from November to March to increase the likelihood of infection with other respiratory viruses, median age 43 [26-56], 50.0% females)(*68*), samples from patients with rheumatic diseases (N=359, median age 52 [41-61], 76.0% females), and eight samples from patients with previous seasonal coronavirus infection collected for routine clinical testing at the Regional Hospital Feldkirch. Sera with PCR-confirmed hCoV infection (hCoV 229-E, n=3; hCoV NL63, n=2 [one of which with 229E co-infection], hCoV OC43, n=2; non-typed, n=2) were drawn between January 2019 and February 2020 and were kindly provided by Andreas Leiherer (Vorarlberg Institute for Vascular Investigation and Treatment VIVIT, Dornbirn, AT). A set of 14 sera of the above (not including hCoV sera) was used for pre-validation of SARS-CoV-2 antigens in an ELISA. Samples (except for those from patients after seasonal coronavirus infection) were processed and stored according to standard operating procedures within the MedUni Wien Biobank facility in a certified (ISO 9001:2015) environment (*69*)

##### Pre-COVID Cohort, Austrian Institute of Technology

Control sera from AIT covered 210 samples of blood donors were obtained in 2014 from the Austrian Red Cross blood bank; collected samples have been stored at −80°C without any freeze thaw cycles.

### 5.6 Pre-validation of antigens using seroreactivity assays

#### 5.6.1 Luminex Assay

In-house produced SARS-CoV-2 RBD, tRBD and NP as well as spike proteins of hCoV HKU-1, OC43, NL63 and 229E (all from Sino Biological Inc, Beijing, CN) were separately coupled to MagPlex carboxylated polystyrene microspheres (Luminex Corporation, Austin, TX) according to the manufacturer’s instruction, with the following minor modifications: For coupling, five µg of each antigen was used per one million microspheres. Coupling was performed in a total volume of 500 µL in 96-Well Protein LoBind Deepwell plates (Eppendorf, Hamburg, DE) and plates were incubated at 600 rpm on a Heidolph Titramax 1000 plate shaker (Heidolph, Schwabach, DE). After each incubation step plates were centrifuged at 400 g for one minute. To collect the microspheres at the bottom of the plate, plates were placed on a Magnetic plate separator (Luminex Corporation, Austin, TX) and the supernatant was poured off by inverting the plates.

Coupling was performed in 200 µL coupling buffer (50 mM MES, pH 5.0). Microspheres with coupled proteins were stored in Assay buffer (PBS supplemented with 1% (w/V) BSA, 0.05% (w/V) NaN_3,_ pH 7.4) at a final concentration of 10,000 microspheres per µL at 4°C in the dark. Sera of patients and controls were five-fold diluted in PBS-Triton X-100 buffer (PBS supplemented with 1% (V/V) Triton X-100, 0.05% (w/V) NaN_3,_ pH 7.4) and were further diluted 240-fold with Assay buffer. Coupled microspheres (800 beads per sample) were first equilibrated to room temperature for 30 min. Plates were then vortexed for 30 sec and sonicated for 20 sec using a Transsonic T470/H sonicator (Elma Electronics, Wetzikon, CH). The required amounts (based on multiples of samples to be analyzed) of microspheres (+10% excess) were transferred to 1.5 mL Protein LoBind tubes (Eppendorf, Hamburg, DE) and centrifuged for 3 min at 1,200 g. Microtubes were then placed on a Magneto Dynal magnetic tube separator (Invitrogen, Carlsbad, CA), supernatants were carefully removed and microspheres were resuspended in 200 µL Assay buffer. Different microspheres were then combined in a 50 mL Falcon tube to yield a total of 800 microspheres per coupled antigen in 30 µL assay buffer per single measurement. Thirty µL of the mixed microsphere suspension was then transferred to wells of a clear 96-well microplate (Corning Inc, Corning, NY). Assay plates were placed on the magnetic plate separator and supernatants were poured off by inverting the plates. Fifty µL of sera (1:1,200-diluted) or assay buffer (blank samples) was applied to each well. Assays were incubated for two hours at RT on the plate shaker (600 rpm). Assay plates were placed on the magnetic plate holder and the supernatants were poured off by inverting the plates. Microspheres were washed by removing the magnetic plate holder and the addition of 100 µL Wash buffer (PBS; 0.05% (V/V) Tween-20; 0.05% (w/V) NaN_3_; pH 7.4) per well. After two minutes of incubation at room temperature, plates were again placed on the magnetic plate holder and supernatants were poured off. After three wash steps 50 µL of a 1:1 mixture of 2.5 μg/mL goat anti-human R-Phyco AffiniPure F(ab’)_2_, Fcγ-specific (# 109-116-098) and F(ab’)_2_-specific IgG (# 109-116-097, both Jackson ImmunoResearch Laboratories Inc., West Grove, PA) in Assay buffer were added. Plates were incubated for 1 h at room temperature on the plate shaker (600 rpm) in the dark. Microspheres were then washed again three times and microspheres were resuspended in 100 µL Assay buffer and median fluorescence intensity (MFI) was immediately measured on a Flexmap 3D Suspension Array System (BioRad, Hercules, CA) with a minimal Count of 100 per microsphere type, a DD Gating of 7,500-25,000 and the Reporter Gain set to “Enhanced PMT (high)”. MFI values were extracted from FM3D result files. A minimum microsphere count of 25 counts was set as cut-off. All samples and single bead types analysed fulfilled the minimum bead count criterium. FM3D results files were compiled in Microsoft Excel and were log2-transformed and blank-corrected by subtracting the mean MFI values of blank samples (assay buffer only) from MFI values of the test samples.

#### 5.6.2 ELISA Assay

Initially, ELISA conditions were optimized in terms of antigen coating conditions (0.5 – 8 µg/mL) and serum-dilutions (1:50 – 1:3,200) to optimize the tradeoff between background seroreactivity and sensitivity in samples from individuals with weak antibody responses. The final protocol was as follows: SARS CoV-2 and hCoV antigens (see above) were diluted to 6 µg/mL in phosphate-buffered saline (PAN Biotech #P-04-36500) and 50 µL were added to each well of MaxiSorp 96-well plates (Thermo #442404). After incubation at 4°C overnight, wells were washed 3x with PBS + 0.1% Tween-20 (Merck #8.22184, PBS-T) and blocked for 1 hour at room temperature with PBS-T + 3% (w/V) milk powder (Fluka #70166). Serum samples were diluted 1:200 in PBS-T + 1% (w/V) milk powder. 100 µL were applied to each well and plates were incubated for 2 h at RT with shaking (450 rpm). Plates were washed 4x before incubation with goat anti-human IgG (Fc-specific) horseradish peroxidase (HRP) conjugated antibodies (Sigma #A0170; 1:50,000 in PBS-T + 1% (w/V) milk powder, 50 µL/well) for 1 h at RT while shaking. After 4 washes, freshly prepared substrate solution (substrate buffer [10 mM sodium acetate in dH_2_O, pH 5, adjusted with citric acid] + 1:60 TMB-stock [0.4 % Tetramethylbenzidine (Fluka #87748) in DMSO] + 1:300 H_2_O_2_ [0.6% in dH_2_O) was applied (150 µL/well) and plates were incubated for 25 minutes at RT with shaking. Reactions were stopped by the addition of 1 M sulfuric acid (25 µL/well). Absorbance was measured at 450 nm on a Tecan Sunrise Microplate reader using a reference wavelength of 620 nm and the Magellan V 7.2 SP1 Software.

### 5.7 TECHNOZYM Anti-SARS-CoV-2 RBD and NP IgG ELISAs Assays

The above-described methodology was slightly adapted for the development of the TECHNOZYM Anti SARS-CoV-2 NP and RBD IgG ELISA test kits (Technoclone, Vienna, AT). The tests plates were provided with the antigens coated at a concentration of 6 µg/mL and lyophilized according to a proprietary in-house protocol. The RBD test kit employs the described tRBD as coating antigen. To allow for a quantitative measurement of SARS-CoV-2 antibody levels, a calibrator set consisting of five calibrators with assigned values was provided for the creation of a calibration curve and was run in parallel with the patients’ samples. The calibrated values were established using the monoclonal antibody CR3022 as a reference material, with 1 U equivalent to 100 ng/mL mAb CR3022 (#Ab01680-10.0, Absolute Antibody, Oxford, UK). The calibrator set covered the concentration range 0 – 100 U/mL and concentrations of anti SARS-CoV-2 IgG antibodies recognizing either tRBD or NP in patient sera could be read directly from the calibration curve.

### 5.8 Technozym NP and RBD IgG ELISA Test validations

The established NP and RBD IgG ELISA assays were either processed manually and analyzed on a Filtermax F5 plate reader (Molecular Devices, San José, USA) or on an Immunomat instrument (Serion Diagnostics, Würzburg, DE) according to the manufacturer’s instructions. IgG antibody levels were reported as numeric values in form of arbitrary U/mL derived from the five-point calibration curve. Cut-offs for test validations were determined by ROC-analysis and the non-parametric 99th right-sided percentile method (CLSI C28-A3). Sensitivities, specificities, PPV, and negative predictive values (NPV, both at 5% estimated seroprevalence) were calculated. ROC-analysis data from automated tests (including Abbott ARCHITECT SARS-CoV-2 IgG, DiaSorin LIAISON^®^ Anti-SARS-CoV-2 S1/S2 IgG) were available for 64 of the positive and 1117 of the negative samples from a previously published study (*30*).

### 5.9 SARS-CoV-2 Neutralisation Assay

A tissue culture infectious dose (TCID_50_) assay for authentic SARS-CoV-2 virus was developed for the determination of neutralizing antibodies. The virus was originally isolated from a clinical specimen, a nasopharyngeal swab taken in mid-March 2020 from a 25-year old male patient in Lower Austria, and was further passaged twice on Vero E6 TMPRSS-2 cells in Dulbecco’s modified Eagle’s medium (DMEM) with 10% (V/V) fetal bovine serum (FBS). Vero E6 TMPRSS-2 cells, initially described in Hoffmann *et al*.(*70*) were kindly provided by Stefan Pöhlmann; Deutsches Primatenzentrum, Göttingen, Germany.

Briefly, assays were performed with Vero 76 clone E6 cells (CCLV-RIE929, Friedrich-Loeffler-Institute, Riems, Germany) cultured in minimum essential medium Eagle (E-MEM) with BioWhittaker Hank’s balanced salt solution (HBSS) (Lonza, Basel, CH) supplemented with 10% (V/V) FBS (Corning Inc, Corning, NY). Neutralizing antibody titers in human serum and plasma were determined as previously described (*71*) with the following alterations: the heat-treated sera were diluted 1:4 in triplicates in serum-free HEPES-buffered DMEM medium. In the case neutralizing antibody titers were determined in human lithium heparin plasma, no heat-treatment was applied and the medium was supplemented with 1x Antibiotic/Antimycotic solution (Thermo Fisher Scientific, Waltham, MA). The heat treatment had no effect on neutralizing titers, as verified in a pre-experiment on SARS-CoV-2 positive and negative plasma samples. In addition, a toxicity control, which was processed the same way as plasma samples, was included. Here, no virus was added, to prevent a false readout of the assay. Cytopathic effect (CPE) was evaluated and scored for each well using an inverted optical microscope. To determine neutralization titer the reciprocal of the highest serum dilution that protected more than 50% of the cells from the CPE was used and was calculated according to Reed and Muench (*72*) .Briefly, assays were performed with Vero 76 clone E6 cells (CCLV-RIE929, Friedrich-Loeffler-Institute, Riems, Germany) cultured in minimum essential medium Eagle (E-MEM) with Hank’s balanced salt solution (HBSS) (BioWhittaker, Lonza, Szabo Scandic, Austria) supplemented with 10% (v/v) fetal bovine serum (Corning, Szabo Scandic, Austria) (FBS). For the assay both sera and plasma was used.

### 5.10 Statistical analyses

Raw data were assessed for normality of distribution and homogeneity of variances using the D’Agostino–Pearson omnibus test before statistical procedures. Differences in median seroreactivities between pre-COVID and COVID sera were compared using the Mann-Whitney U tests on blank-corrected log2-transformed median fluorescence intensities (Luminex data) or OD_490_ absorbances (ELISA), respectively. Correlation analyses of nonparametric data were performed by Spearman’s rank-order correlation (r_s_), otherwise Pearsons’ correlation (r) was used. Relative IgG signals of outliers against SARS-CoV-2 and hCoV antigens were compared by One-Way ANOVA followed by a Sidak test to correct for multiple comparisons. ROC-analysis data from automated tests were compared to the established ELISA tests according to DeLong. Sensitivities and specificities were compared by z-tests. Data on the diagnostic performances of antigens and cross-reactivity were analyzed using Graphpad Prism Version 8.1.0 (GraphPad Software, San Diego, CA, USA) Validation data were analyzed using MedCalc v19 (MedCalc Software, Ostend, Belgium) and Analyse-it 5.66 (Analyse-it Software, Leeds, UK) and SPSS 23.0 (SPSS Inc.). Data from SARS-CoV-2 acute sera from hospitalized individuals or outpatients obtained by the B&S Central Laboratory Linz were statistically analyzed with the MedCalc 13.1.2.0.

## Data Availability

Data and study protocol can be requested from the corresponding authors by interested researchers.

## Acknowledgements

We thank Florian Krammer (Icahn School of Medicine at Mount Sinai, NY, United States) for providing the constructs used for the production of RBD and CR3022. We thank George Lomonossoff (John Innes Centre, Norwich, UK) and Plant Bioscience Limited (PBL) (Norwich, UK) for supplying the pEAQ-HT expression vector. The authors thank Naila Avdic, Kristina Jagersberger, Christina Hausjell, Viktoria Mayer, Anna-Carina Frank, Sophie Vazulka, Florian Mayer, Matthias Müller, Andreas Dietrich, Mathias Fink, Florian Weiss, Giulia Borsi, Mohammed Hussein, Patrick Mairhofer, Andreas Fischer, Alexander Doleschal (all Department of Biotechnology, University of Natural Resources and Life Sciences (BOKU) Vienna or and/or ACIB GmbH affiliated) for their support in the cloning, production and analysis of SARS-CoV-2 antigens described in this work. The BLI and mass spectrometry equipment was kindly provided by the EQ-BOKU VIBT GmbH and the BOKU Core Facilities *Biomolecular & Cellular Analysis* (BmCA) and *Mass Spectrometry*. Boehringer Ingelheim RCV GmbH & Co KG fully supported the endeavor and granted access to manufacturing technologies for process development and manufacturing.

The authors thank Irene Schaffner and Jakob Wallner (BOKU BmCA) for assisting in BLI measurements and ForteBio for providing SAX biosensors. The authors want to thank Maria Ozsvar-Kozma, Patrick Mucher, Manuela Repl and Astrid Radakovics (Department of Laboratory Medicine, Medical University Vienna) for outstanding technical assistance. The authors thank the following collaborators for providing biomaterial and data for ELISA evaluation: Manfred Nairz, Sabina Sahanic, Thomas Sonnweber, Alex Pizzini, Ivan Tancevski (Department of Internal Medicine II, Medical University of Innsbruck), Markus Anliker, Lorin Loacker, Wolfgang Prokop (Central Institute for Medical and Chemical Laboratory Diagnosis, Innsbruck University Hospital), Wolfgang Mayer, Harald Schennach (Central Institute for Blood Transfusion & Immunological Department, University Hospital of Innsbruck), Daniel Aletaha (Department of Internal Medicine III, Medical University of Vienna), Robab Breyer-Kohansal, Otto C Burghuber, and Sylvia Hartl (LEAD-Study - Ludwig Boltzmann Institute for Lung Health, Vienna, Austria), Andreas Leiherer (Vorarlberg Institute for Vascular Investigation and Treatment [VIVIT], Dornbirn, Austria).

Finally, the BOKU Spin-off Novasign GmbH supported the sustainability concept of BOKU by creating and hosting the BOKU-COVID19 Portal (portal.boku-covid19.at) that enables researchers worldwide to obtain trial samples of the within this work described SARS-CoV-2 antigens free of charge. Armin Khodaei and Roger-Dalmau-Diaz implemented the database and interface of the portal.

## Author Contributions

A.J.,A.W.,C.J.B.,E.L.,F.E.,F.G.,G.S.,G.W.K.,H.H.,J.M.,M.S.,M.D.,M.W.,M.K.,M.C.P.,N.L.,N.Bo.,N.Bi.,P.P.A., R.G.,T.P.,W.G. designed study concepts.

A.J.,A.W.,C.J.B,F.E.,F.G.,G.S.,J.M.,M.W.,M.K.,M.C.P.,N.Bi.,R.G.,W.G. acquired financial support for this project.

A.W.,B.H.,B.D.,C.J.B,E.G.,E.L.,F.G.,G.M.,G.S.,G.SM.,G.W.K.,H.H.,J.H.,J.B.,J.M.,Li.M.,M.H.,M.E.,M.S.,M.D., M.W.,M.K.,M.C.P.,N.L.,N.M.,N.Bi.,P.P.A.,P.S.,R.H.,R.G.,Ri.S.,T.P.,W.G. developed methodology.

A.E.E.,A.G.,A.W.,B.H.,B.D.,C.I.,C.J.B,C.K.,C.T.,C.G.G,D.M.,D.S.,E.G.,E.L.,F.S.,G.M.,G.L.,G.SM.,G.W.K.,G.H., J.H.,J.D.V.,J.L.R.,J.B.,Lu.M.,M.H.,M.S.,M.K.B.,M.D.,M.W.,M.K.,N.L.,N.M.,N.Bi.,P.P.A.,P.Q.,P.S.,R.H.,R.G.,Ri. S.,Ro.S.,R.B.W.,T.P.,U.V. conducting a research and investigation process, specifically performing the experiments, or data/evidence collection.

K.V. developed evaluation software code.

A.E.E.,A.G.,B.D.,C.I.,F.G.,G.H.,H.H.,M.D.,M.K.,R.B.W.,T.P. performed data curation.

A.W.,F.G.,H.H.,J.H.,M.H.,M.S.,M.K.,W.G. performed data validation and evaluated data reproducibility.

A.E.E.,A.G.,A.W.,B.H.,B.D.,C.I.,C.G.G,D.M,E.L.,F.G.,G.M.,G.SM.,G.W.K.,G.H.,H.H.,J.D.V.,K.V.,Lu.M.,M.E., M.D.,M.K.,M.C.P.,N.L.,P.P.A.,Ri.S. performed formal analysis to analyze and synthesize data.

H.H.,M.D.,M.K.,R.B.W. performed data visualization.

A.E.E.,A.J.,A.G.,C.I.,C.J.B,D.S.,E.L.,F.E.,F.G.,G.S.,G.L.,G.H.,G.W.,J.D.V.,J.L.R.,J.M.,Lu.M.,M.K.B.,M.W.,M.C.P., N.Bo.,N.Bi.,P.Q.,R.H.,R.G.,Ro.S.,R.B.W.,W.G., provided study materials, reagents, laboratory samples, animals, instrumentation, computing resources, or other analysis tools.

A.W.,C.J.B,F.G.,G.S.,G.W.,Lu.M.,M.C.P,M.D.,M.K.,N.Bo.,N.Bi.,R.G.,R.B.W.,W.G. were responsible for supervising research activities and for project administration.

H.H.,M.D.,M.K.,M.C.P.,R.G.,T.P. wrote the original draft.

B.D.,C.J.B,E.L.,F.G.,G.W.K.,J.D.V.,J.M.,Lu.M.,M.E.,M.C.P.,N.L.,P.P.A.,P.S.,R.G.,W.G. reviewed and edited the manuscript.

## Data and materials availability

All data needed to evaluate the conclusions in the paper are present in the manuscript and/or the Supplementary Materials. Recombinant SARS-CoV-2 antigens can be requested from interested researchers for research purposes under www.boku-covid19.at (after registration). For commercial purposes antigens can be requested from enGenes Biotech GmbH. Technozym NP and RBD IgG serotests are available from Technoclone (Technoclone Herstellung von Diagnostika und Arzneimitteln GmbH, Vienna, Austria)

