## Supplementary Materials for "A comprehensive antigen production and characterization study for easy-to-implement, highly specific and quantitative SARS-CoV-2 antibody assays"

**This PDF file includes:**

Fig. S1. MS spectra of the two RBD glycosites A) SIVRFPNITNLCPFGE and B) VFNATRFASVYAWNRK.
Fig. S2. HP-SEC elution profiles of RBD produced in different expression systems.
Fig. S3. Biolayer-Interferometry analysis of SARS-CoV-2 antigens with specific antibodies.
Fig. S4. Purity of HEK-expressed tRBD is of utmost importance for sensitive anti-RBD IgG detection.
Table S1. Characteristics of SARS-CoV-2 positive samples used for test evaluation.
Fig. S5. ROC curve analysis of the Technozym NP and RBD ELISA in comparison with CE-marked automated Abbott and DiaSorin test systems.
Table S2. Time-resolved monitoring of SARS-CoV-2 NP- and tRBD-specific antibody levels and neutralization titers
Fig. S6. Specificity control testing with RBD variants derived from different expression systems.

peptide: SIVRFPNITNLCPFGE
mass: 1863.9421
range: 1000-2000

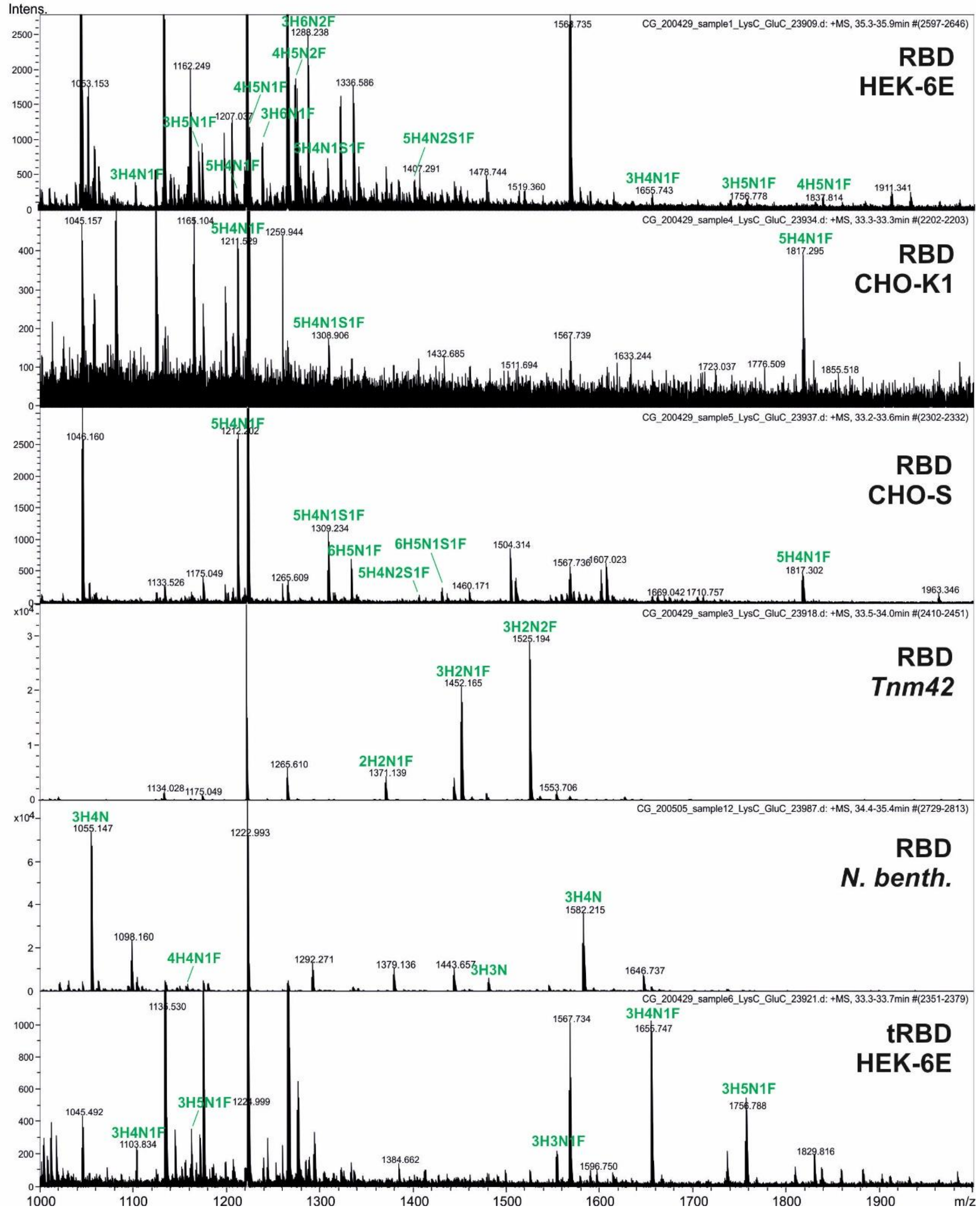

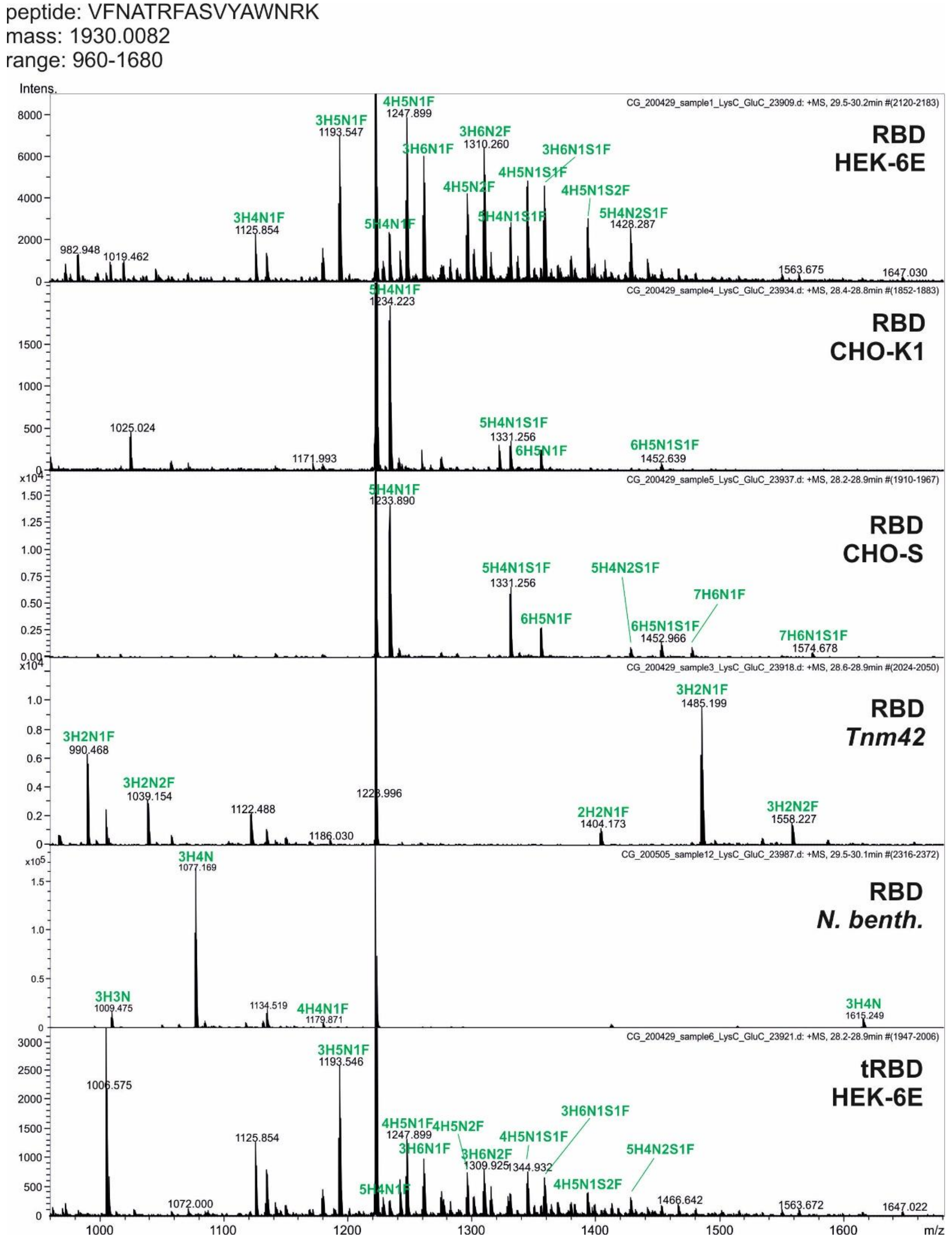


**Fig. S1. MS spectra of the two RBD glycosites A) SIVRFPNITNLCPFGE and B) VFNATRFASVYAWNRK.**

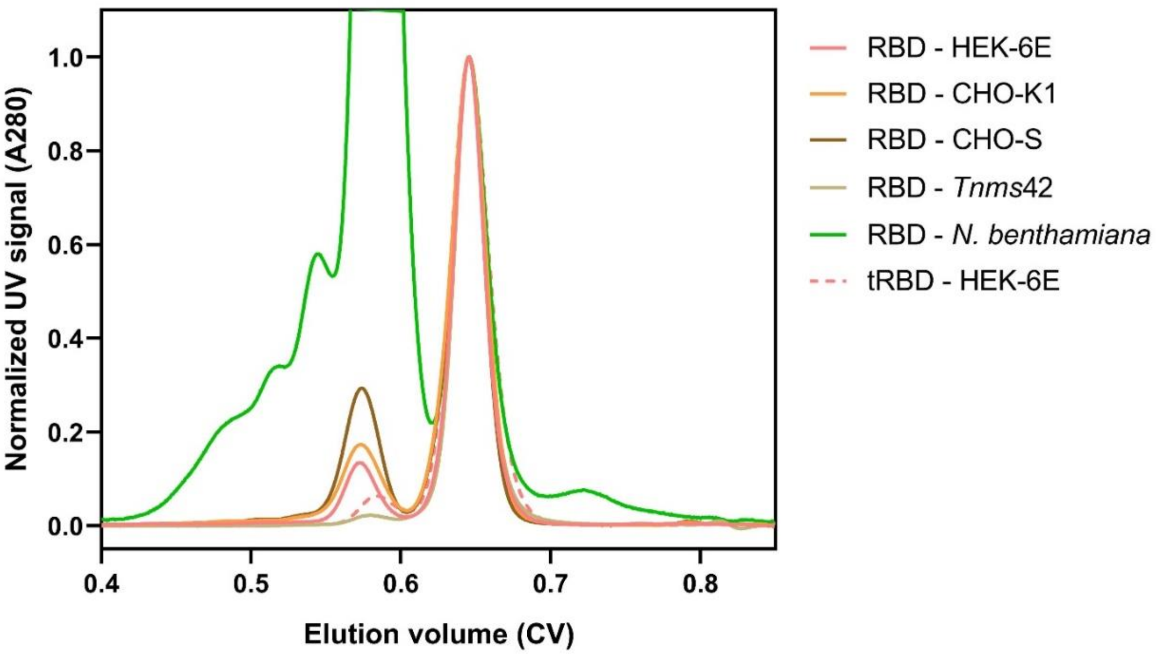


**Fig. S2. HP-SEC elution profiles of RBD produced in different expression systems.** Content of monomer was normalized in order to allow for dimer comparison. All proteins where purified with IMAC and AIEX, except for *Tnms*42 RBD which was purified with IMAC and preparative SEC resulting in dimer removal.

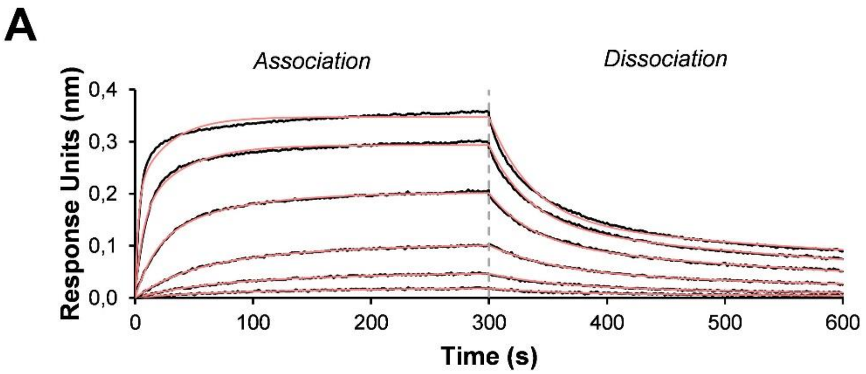


| | $K_D$1 (nM) | $K_D$2 (nM) | $k_a$1 ($M^{-1}$ $s^{-1}$) | $k_a$2 ($M^{-1}$ $s^{-1}$) | $k_d$1 ($s^{-1}$) | $k_d$2 ($s^{-1}$) |
|---|---|---|---|---|---|---|
| Batch 1 | 25 | 8 | 1.1E+06 | 2.7E+05 | 2.7E-02 | 1.8E-03 |
| SEM | 1.4 | 0.7 | 5.8E+04 | 2.1E+04 | 5.3E-04 | 8.4E-05 |
| Batch 2 | 28 | 8 | 9.2E+05 | 3.7E+05 | 2.5E-02 | 2.4E-03 |
| SEM | 1.6 | 0.9 | 3.1E+04 | 1.7E+04 | 6.6E-04 | 2.7E-04 |

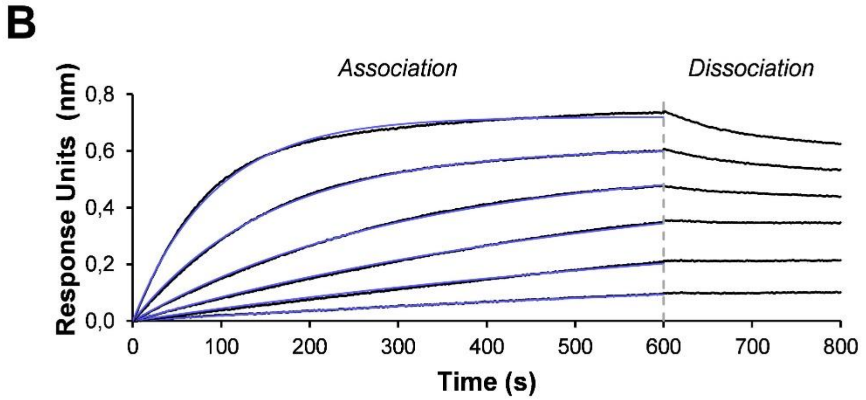


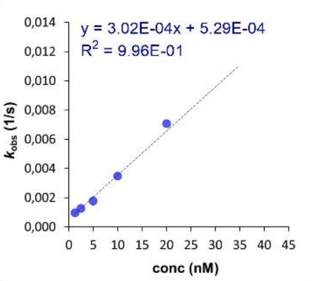


| Conc. (nM) | $k_{obs}$(1/s) | SEM |
|---|---|---|
| 1.25 | 5.1E-04 | 2.1E-04 |
| 2.5 | 6.5E-04 | 1.2E-04 |
| 5 | 1.5E-03 | 7.8E-05 |
| 10 | 3.4E-03 | 9.6E-05 |
| 20 | 6.9E-03 | 2.1E-04 |
| 40 | 1.2E-02 | 5.0E-04 |

**Fig. S3. Biolayer-Interferometry analysis of SARS-CoV-2 antigens with specific antibodies**.

**A)** Binding kinetics of the interaction between biotinylated mAb CR3022 loaded on SAX biosensors and tRBD at a concentration range of 1.2 - 300 nM. Representative real-time association and dissociation curves are shown. The lower panel gives the mean kinetic parameters of a quadruplicate measurement from two independent tRBD production batches. B) Representative binding curves of the interaction of a biotinylated commercially available anti-NP mAb (ab272852, Abcam) and NP (1.25 – 40 nM). As kinetic parameters could not be calculated, a surrogate kinetic parameter, $k_{obs}$, is given as a mean of two duplicate measurements of two independently produced NP batches in the lower panel. Black lines represent the response curves of the association and dissociation. Fitted curves are shown as red and blue lines, respectively, dashed vertical lines indicate the transition between association and disassociation phases. $K_D$, equilibrium dissociation constant; $k_a$, association rate constant; $k_{d,}$ dissociation rate constant, $k_{obs}$, observed binding rate constant, SEM, standard error of the mean, Conc., concentration

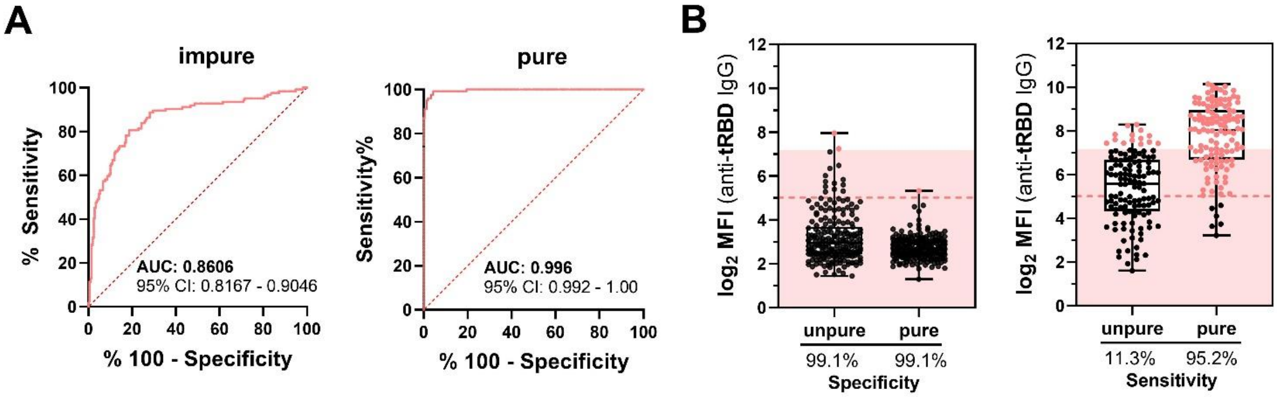


**Fig. S4. Purity of HEK-expressed tRBD is of utmost importance for sensitive anti-RBD IgG detection.** HEK-expressed tRBD was either only IMAC-purified (impure, 87.5% purity, as determined by HP-SEC) or was additionally polished using anion-exchange chromatography. Pure tRBD gives the blank-corrected mean of three individual production batches (mean purity: 97.5%). Seroreactivity with convalescent COVID-sera (n=110) and 210 pre-COVID sera was assessed by a Luminex assay. **A)** ROC curves with AUC-analysis for both proteins, also indicating the 95% confidence intervals. **B)** Seroreactivity of individual sera at a serum dilution of 1:1,200. Cut-offs were pre-defined to ensure a specificity of 99.1%. Shades indicate the cut-off for impure tRBD, dashed lines indicate the cut-off for pure tRBD (both calculated from the ROC curve). Pink circles among the pre-COVID sera indicate sera that are above the cut-off, while black circles among the COVID-sera indicate sera that fall below the cut-off.

| Collection site | N | Age [y] | Female sex | Median days after PCR/<br>symptom onset* | Severity |
|---|---|---|---|---|---|
| Vienna | 70 | 49 (37 – 56) | 34 (49%) | 43 (IQR 28 – 51) | asymptomatic=5 (7%)<br>mild=29 (41%)<br>moderate=18 (26%)<br>severe=4 (6%)<br>hospitalized=14 (20%) |
| Innsbruck | 174 | 54 (39 – 62) | 113 (65%) | 54 (IQR 45 – 65) | outpatient=71 (41%)<br>hospitalized, general ward= 75 (43%)<br>Hospitalized, intensive care = 28 (16%) |

**Table. S1. Characteristics of SARS-CoV-2 positive samples used for test evaluation.**

*symptom onset for symptomatic patients/donors from Vienna, first positive PCR for asymptomatic donors from Vienna and patients/donors from Innsbruck.

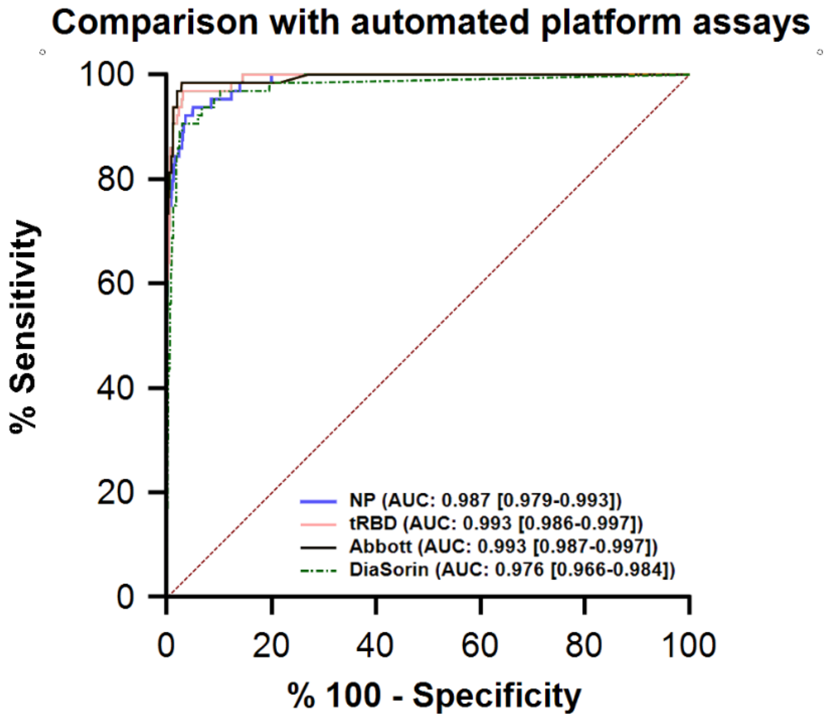


**Fig. S5**. **ROC curve analysis of the Technozym NP and RBD ELISA in comparison with CE-marked automated Abbott and DiaSorin test systems**. Test performance was validated with the MedUni Wien Biobank pre-COVID cohorts (N=1,117, excluding the described 8 hCoV samples) and COVID cohorts (N=64).

| **Symptom onset (days)** | **n (%)** | **NT assay (titer)** | **TC NP IgG (U/mL)** n negative (%) n positive (%) | **TC RBD IgG (U/mL)** n negative (%) n positive (%) |
|---|---|---|---|---|
| **1-5 (days)** | **34 (100%)** | | **29 neg. (85.3%)** | **32 neg. (94.1)** |
| median | | N/A | 0.6 | 0.3 |
| range | | N/A | (0.0-3.5) | (0.0-3.9) |
| IQR | | N/A | (0.2-1.3) | (0.1-0.6) |
| | | | **5 pos. (14.7%)** | **2 pos. (5.9%)** |
| median | | 1:16 | 21.3 | 8.8 |
| range | | (<1:4-1:128) | (5.5-76.0) | (7.7-9.9) |
| IQR | | (1:5-1:48) | (6.6-59.5) | N/A |
| **6-10 (days)** | **35 (100%)** | | **19 neg. (54.3%)** | **23 neg. (65.7%)** |
| median | | N/A | 0.6 | 0.4 |
| range | | N/A | (0.0-3.7) | (0.0-2.7) |
| IQR | | N/A | (0.2-1.3) | (0.2-1.0) |
| | | | **16 pos. (45.7%)** | **12 pos. (34.2%)** |
| median | | 1:48 | 32.0 | 17.6 |
| range | | (1:4->1:512) | (6.0-166) | (5.0-121.0) |
| IQR | | (1:32-1:172) | (10.7-95.8) | (7.6-50.0) |
| **11-15 (days)** | **17 (100%)** | | **4 neg. (23.5%)** | **6 neg. (35.3%)** |
| median | | N/A | 0.0 | 0.1 |
| range | | N/A | (0.0-0.1) | (0.0-3.3) |
| IQR | | N/A | (0.0-0.1) | (0.0-2.2) |
| | | | **13 pos. (76.5%)** | **11 pos. (64.7%)** |
| median | | 1:192 | 78.6 | 133.8 |
| range | | (<1:4->1:512) | (24.6-1732.4) | (5.2-328.8) |
| IQR | | (1:48-1:1,024) | (51.0-126.4) | (98.3-206.4) |
| **16-22 (days)** | **18 (100%)** | | **0 neg. (0%)** | **0 neg. (0%)** |
| median | | N/A | N/A | N/A |
| range | | N/A | N/A | N/A |
| IQR | | N/A | N/A | N/A |
| | | | **18 pos. (100%)** | **18 pos. (100%)** |
| median | | 1:320 | 202.6 | 146.6 |
| range | | (1:32->1:512) | (30.9-4064.0) | (5.1-447.6) |
| IQR | | (1:128-1:1,024) | (109.0-933.8) | (61.3-217.5) |

[a] NT assay: negative (titer <1:4); positive (titer ≥1:4)

[b]TC NP IgG ELISA: negative (<5.000 U/mL); positive (≥5.000 U/mL)

[c]TC RBD IgG ELISA: negative (<5.000 U/mL); positive (≥5.000 U/mL)

**Table S2**. **Time-resolved monitoring of SARS-CoV-2 NP- and tRBD-specific antibody levels and neutralization titers**. Titers for the SARS-CoV-2 NT assay, and Units/mL (U/mL) for the TC NP and RBD IgG ELISAs in 64 patients with SARS-CoV-2 RT-PCR confirmed COVID-19 with serial blood samples (n=104) collected at different time points from symptom onset. For descriptive statistics and statistical analyses a neutralization titer below 1:4 was assigned a value of 1:2 and a titer above 1:512 was assigned 1:1,024. Likewise, RBD and NP ELISA titers of 0 U/mL were assigned a value of 0.1 U/mL.

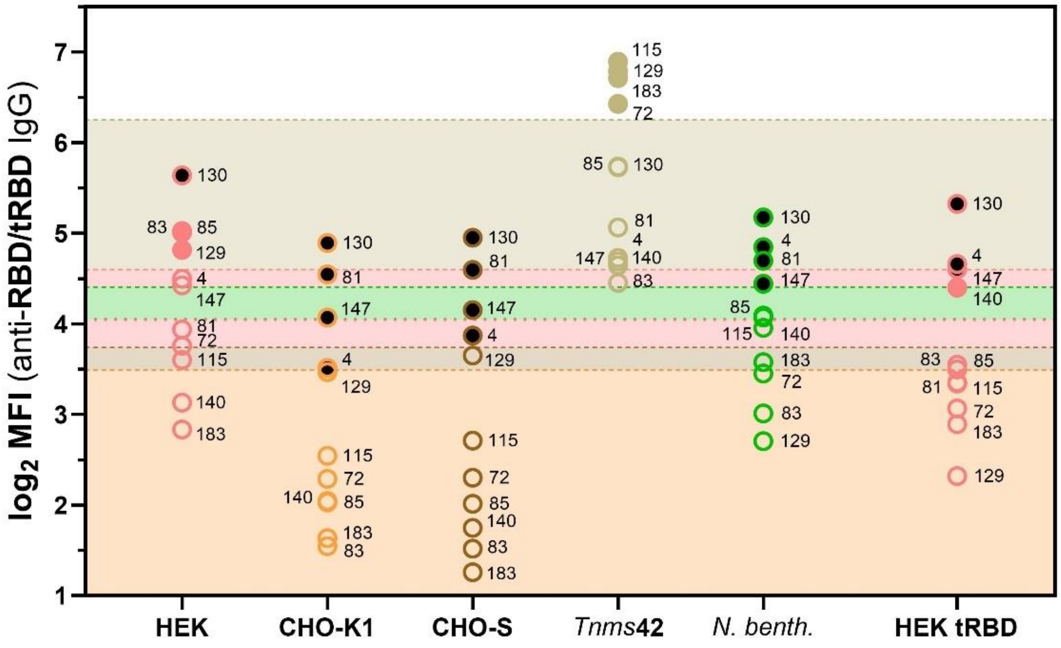


**Fig. S6. Specificity control testing with RBD variants derived from different expression systems.** Sera (serum dilution: 1:1,200) were from a pre-COVID cohort (n=210) and were identified as false-negative using the Luminex platform with a pre-specified specificity of 99.1%. Numbers represent the identification codes of the respective sera. Shades and dashed lines indicate the respective cut-offs color-coded according to the different expression systems. Open circles indicate sera that fall below the cut-off. Full circles indicate outlier sera that fall above the cut-off and are considered as false-positive; black-filled circles indicate sera that are identified as false-positive with antigens from at least three expression platforms.
